# Extreme precipitation, exacerbated by anthropogenic climate change, drove Peru’s record-breaking 2023 dengue outbreak

**DOI:** 10.1101/2024.10.23.24309838

**Authors:** Mallory J. Harris, Jared T. Trok, Kevin S. Martel, Mercy J. Borbor Cordova, Noah S. Diffenbaugh, César V. Munayco, Andrés G. Lescano, Erin A. Mordecai

## Abstract

**Highlights:**

- In March 2023, Cyclone Yaku was followed by a large dengue epidemic in northwest Peru
- Extreme precipitation during Cyclone Yaku caused 60% of dengue cases
- More cyclone-attributable cases occurred in warm, urban, flood-susceptible districts
- Global warming has increased the risk of warm, very wet March weather in the region

**Science for Society:** Anthropogenic climate change is increasing the risk of extreme weather that can lead to infectious disease epidemics, but few studies have directly measured this health consequence of climate change. Extreme precipitation can drive outbreaks of mosquito-borne diseases by displacing people, disrupting public health activities, and creating aquatic breeding habitat. Here, we quantify the effects of extreme precipitation during Cyclone Yaku in northwestern Peru in March 2023. The cyclone was immediately followed by a dengue outbreak where cases exceeded historic averages by tenfold. We estimate that 60% of cases (or 22,014 cases) reported over three months in the affected districts were attributable to extreme precipitation during Cyclone Yaku. Compared with the preindustrial era, extremely wet March conditions were 31% more likely to occur (and 189% more likely to co-occur with warm temperatures suitable for dengue transmission) in recent decades in northwestern Peru. Assessing the linkages between climate change, extreme weather, and outbreaks of dengue and other infectious diseases is crucial for understanding the current impacts of climate change and for preparing for future health risks.

**eTOC Summary:** This study examines relationships between historical climate forcing, extreme weather, and health, focusing on Cyclone Yaku and Peru’s 2023 dengue epidemic. Historical climate forcing has increased the likelihood of extreme precipitation coinciding with warm temperatures suitable for transmission in March in northwest Peru. In turn, extreme precipitation during Cyclone Yaku caused a majority of dengue cases in the epidemic, especially across warmer districts. Extreme weather, made more likely by climate change, is already having an impact on human health.

Climate change is increasing the likelihood of extreme weather that can drive outbreaks of climate-sensitive diseases. For example, dengue burden has recently increased rapidly with unusually warm and wet conditions. However, linkages between historical climate change, extreme weather, and mosquito-borne disease have not been traced quantitatively. Here, we analyze the contribution of extreme precipitation to dengue in northwestern Peru during Cyclone Yaku in March 2023. Using generalized synthetic control methods, we estimate 60% of cases were attributable to extreme precipitation and more cyclone-attributable dengue cases occurred in warmer, more flood susceptible, and more urban districts. Historical climate forcing has increased the likelihood of concurrent extreme precipitation and warm temperatures suitable for dengue transmission in northwestern Peru in March by 189%. This case study is one of the first to estimate cases of mosquito-borne illness caused by extreme weather conditions and shows those conditions were made more likely by climate change.

## 1 Introduction

Human activities are driving major changes to the climate system, including more frequent and intense extreme weather events such as heat waves, droughts, and cyclones (Seneviratne et al. 2021; National Academies of Sciences, Engineering, and Medicine 2016). An estimated net of 60,000 deaths and $260B in economic damages have been attributed to extreme weather linked to climate change (Newman and Noy 2023). Prior work found that climate change has increased the risk of an array of adverse health outcomes, including heat-related deaths, Lyme disease, and foodborne illness connected to the bacteria *Vibrio vulnificus* (National Academies of Sciences, Engineering, and Medicine 2016; Ebi et al. 2020; Hegerl et al. 2010; Vicedo-Cabrera et al. 2021; Chapman et al. 2022; Ogden et al. 2014; McPherson et al. 2017; Vezzulli et al. 2016). However, a critical gap remains in measuring the contribution of changes in extreme weather to mosquito-borne disease outbreaks (Carlson et al. 2024). Although mosquito-borne disease transmission is known to be sensitive to climate, including extreme weather (Shocket et al. 2020; Mordecai et al. 2019; Alcayna et al. 2022; Mora et al. 2022), few studies have attempted to quantify the contribution of historical climate change to mosquito-borne disease burden (e.g., (Carlson et al. 2023; Childs et al. 2024)), and these studies have focused on the contribution of long-term increases in mean temperature.

Mosquito-borne diseases present a large and growing health threat, causing over 650,000 deaths annually (World Health Organization 2024). Reported cases of dengue, a virus transmitted by *Aedes aegypti* and *Ae. albopictus*, have surged by a factor of more than twenty in recent years compared to 2000, with a total of over five million cases and 5,000 associated deaths reported globally in 2023 (World Health Organization 2023). Some have speculated that extreme weather (particularly extreme precipitation) is a primary driver of this increase in transmission (World Health Organization 2023), given that mosquito-borne disease risk is associated with cyclone exposure and heavy rainfall, particularly in dry regions (Shocket et al. 2020; Gibb et al. 2023; Lowe et al. 2021, 2018; Li et al. 2022). However, isolating the causal effects of particular weather conditions requires accounting for multiple time-varying drivers that influence dengue transmission, which are often inadequately captured by existing data sources and modeling approaches (e.g., strain introductions, immunity, mobility, vector control, time spent outdoors by humans, personal protective behavior, vector competence and abundance, and urbanization) (Zhang et al. 2020; Nova et al. 2021; Giesen et al. 2020; Ogden 2017).

Additional climate and socioeconomic factors may modulate the effects of extreme weather on dengue incidence. For example, the built environment (e.g., greater urban infrastructure, reduced water access, housing built from low-quality materials) and human behavior (e.g., water storage practices) may increase vulnerability to dengue outbreaks after extreme weather (Rufasto Goche et al. 2025; Rahman et al. 2021; Kolimenakis et al. 2021; Nakhapakorn and Tripathi 2005; Piaggio et al. 2024; Gibb et al. 2023; Mulligan et al. 2015; Alcayna et al. 2022; Reiter et al. 2003). The relationship between precipitation and dengue risk is also nonlinear and time-dependent: flooding may promote vector-borne disease transmission by disrupting critical infrastructure and creating mosquito breeding habitat, but can also reduce transmission by flushing out vector habitat (Koenraadt and Harrington 2008, Seidahmed and Eltahir 2016; Lowe et al 2021, 2018; Caldwell et al. 2021). Temperature may also modulate the effects of extreme precipitation on outbreak potential, given known temperature-sensitivity of mosquitoes, with the greatest risk of dengue transmission by *Ae. aegypti* at 29◦C (Mordecai et al. 2019, Caldwell et al. 2021, Mordecai et al 2017). Identifying factors associated with greater risk of large dengue outbreaks following extreme weather events may help guide vector control and public health emergency preparedness (Udayanga et al. 2020).

Finally, quantifying the extent to which historical climate forcing has influenced the likelihood of climate conditions associated with large dengue outbreaks is an important step toward tracing potential linkages between climate change and human health. Attribution studies have quantified the contribution of anthropogenic forcing to particular extreme weather conditions, including heat waves, heavy rainfall, hurricanes, and droughts (Trok et al. 2024; Diffenbaugh et al. 2015; Van Oldenborgh et al. 2017; Diffenbaugh et al. 2017; Emanuel 2017). Although most places are likely to experience increases in the intensity of extreme precipitation at higher levels of climate forcing, the influence of historical climate change on the likelihood of extreme precipitation events varies between regions, underscoring the importance of tailoring attribution analyses to specific geographic areas (Chand et al. 2022; Reed et al. 2022; Murakami et al. 2017; Seneviratne et al. 2021; Diffenbaugh et al. 2017). Given that temperature and precipitation are interdependent and are both changing in response to climate forcing, analyses that examine simultaneous changes in these weather variables are necessary to build a more comprehensive understanding of the impacts of climate change (Diffenbaugh et al. 2015).

Peru is a particularly relevant location for estimating the effect of extreme weather on dengue transmission. In March 2023, the normally dry northwestern coast experienced anomalously heavy precipitation associated with Cyclone Yaku and a coastal El Niño event which caused significant flooding, (Peng et al. 2024; Munayco et al. 2024). Shortly thereafter, the region experienced Peru’s largest recorded dengue outbreak, with case incidence by July 2023 exceeding the five-year average by a factor of ten (6.55 versus 0.64 cases per thousand) and 381 dengue-related deaths reported (Munayco et al. 2024). It has been postulated that this outbreak was linked to extreme precipitation during Cyclone Yaku and the resulting disruption of infrastructure and health systems (Bagcchi 2023; Cabezas Sánchez 2023; Peng et al. 2024), although additional potential contributors include the spread of the DENV-2 serotype throughout the region beginning in 2019 (Bailon et al. 2024) and broader trends related to human mobility, urbanization, and political and financial instability (Munayco et al. 2024; World Health Organization 2023; Cabezas Sánchez 2023). To date, no causal analysis has quantified the specific contribution of extreme precipitation during Cyclone Yaku to the dengue outbreak, limiting understanding of the event’s direct health impact.

Here, we describe linkages between historical climate change, extreme precipitation, and the 2023 dengue outbreak in northwestern Peru. First, we constructed a generalized synthetic control model to estimate how many additional dengue cases were caused by extreme precipitation during Cyclone Yaku, while controlling for potential confounding variables (Xu 2017; Bruhn et al. 2017; Shioda et al. 2021; Nyathi et al. 2019; Sheridan et al. 2022; Schwarz et al. 2023). We found that 60% (95% CI: 24% - 86%) of cases in affected districts were attributable to extreme precipitation, which significantly increases cases for almost three months, causing 22,014 (95% CI: 8,665 – 31,712) out of 36,709 cases. Next, we found that the greatest impacts of extreme precipitation on dengue burden occurred in districts with warmer temperatures during Cyclone Yaku, greater flood susceptibility, and more urban infrastructure. Finally, we analyzed an ensemble of climate model simulations to test whether historical climate change has increased the risk of weather conditions associated with greater dengue transmission in northwestern Peru. We found that historical climate forcing has increased the likelihood of extreme monthly March precipitation in northwestern Peru by 31% in recent decades (1965 – 2014) compared to a preindustrial baseline, while the likelihood of concurrent extreme precipitation and warm temperatures suitable for dengue transmission has increased by 189%. These analyses may inform context-specific public health and risk reduction measures for dengue and other weather-sensitive diseases, in addition to providing a template for understanding the relationships between climate forcing, extreme weather, and human health more broadly.

## 2 Results

### 2.1 Cyclone Yaku caused the most extreme precipitation in districts with warm, dry climates

Prior to conducting causal inference, we first identified the districts with the most extreme precipitation to be compared with a control pool of districts with non-extreme precipitation that are otherwise similar. We identified 49 districts with extreme precipitation during Cyclone Yaku (defined as precipitation anomalies exceeding 8.5 mm/day), which generally had warm and dry climates and aligned with reports of regions impacted by Cyclone Yaku (Table S1, Figure 1B, Figure S1) (SENAMHI 2023; MapAction). There were 474 districts with non-extreme precipitation (precipitation anomalies below 7.0 mm/day) that reported dengue cases in 2023, which we considered for inclusion in the control pool (Figure S2). To construct our control pool, which we used to estimate a counterfactual for the cyclone-affected districts, we used matching to select a set of districts that, on average, more closely reflected the relationship between temperature and precipitation in the districts affected by extreme precipitation during Cyclone Yaku (Figure S3). Following matching, 123 districts, predominantly in the northern coast and across the Amazon basin, were included in the control pool (Figure S2). Matching eliminated periods of especially large imbalance in precipitation between the groups of districts with extreme precipitation and the control pool of districts with non-extreme precipitation, although there was some remaining imbalance where the matched control group tended to be warmer and wetter than the districts that experienced extreme precipitation during Cyclone Yaku (Figure S4, Figure S5, Table S2, see discussion in subsection S1.2).

**Figure 1:**
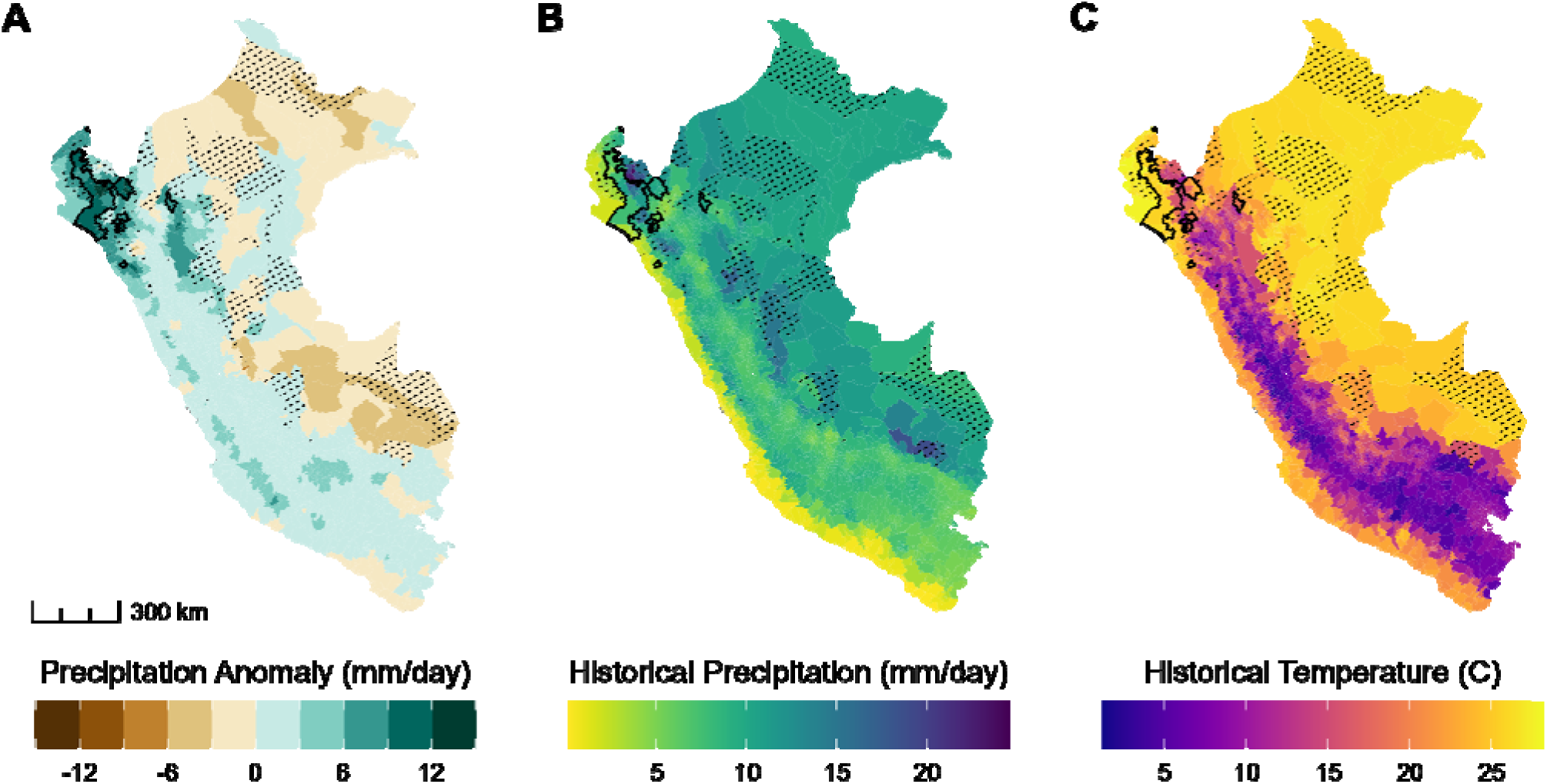
Historical climate conditions across districts in Peru compared to during Cyclone Yaku. In all panels, the bold outline encompasses districts with extreme precipitation during Cyclone Yaku and diagonal dashed lines indicate matched control districts with non-extreme precipitation. (A) Precipitation anomalies (mm/day), calculated as the difference between the mean daily precipitation between March 7 - 20, 2023 and the mean daily precipitation during the same time period across the reference years 1973 - 2022. (B) Average daily precipitation (mm/day) across the historical reference period. (C) Average temperature (in degrees Celsius) across the historical reference period. A scale bar is provided in the bottom left corner.

### 2.2 Sixty percent of dengue cases were attributable to extreme precipitation in the affected districts

Dengue incidence was substantially elevated in all districts following Cyclone Yaku, but particularly across the districts that experienced extreme precipitation during the cyclone (Figure S6). In the districts that experienced extreme precipitation, there were 2.68 cases per thousand people in 2023 compared to the prior monthly average of 0.03 cases per thousand people from 2010 - 2022. The 2023 outbreak peaked at 2.68 cases per thousand people, exceeding the peaks of prior large outbreaks in 2015, 2017, and 2022 (0.46, 0.62, and 0.22 cases per thousand people, respectively) by a factor of more than four. The outbreak in 2017 followed another period of elevated precipitation (Figure S6).

We used a quasi-experimental generalized synthetic control approach to estimate the number of dengue cases attributable to extreme precipitation by comparing observed cases in districts with extreme precipitation to a counterfactual estimate of cases in those districts had they not experienced extreme precipitation, determined based on a pool of control districts (Figure S5, Figure S6). Compared to the synthetic control, districts with extreme precipitation had significantly elevated dengue cases for close to three months (April 22nd - July 14th). During that time period, 22,014 (95% CI: 8,665 - 31,712) out of 36,709 total cases, or 60% (95% CI: 24% - 86%) of cases in affected districts were attributable to extreme precipitation during Cyclone Yaku. Attributable cases peaked between May 20th and June 16th at 9,918 (95% CI: 4,434 - 14,191) cases, constituting 63% (95% CI: 28% - 91%) of reported cases during that time period (Figure 2B, Table S4). Overall, the generalized synthetic control model fit observed cases moderately well prior to the cyclone (unadjusted *R^2^* = 0.67) (Figure 2A), including during the large outbreak in 2017, supporting its use as a counterfactual. Models with five latent factors minimized prediction error during cross-validation (Figure S7; see Figure S8 for corresponding factor loadings). There was a significant positive association between temperature and dengue incidence (Table S3).

**Figure 2:**
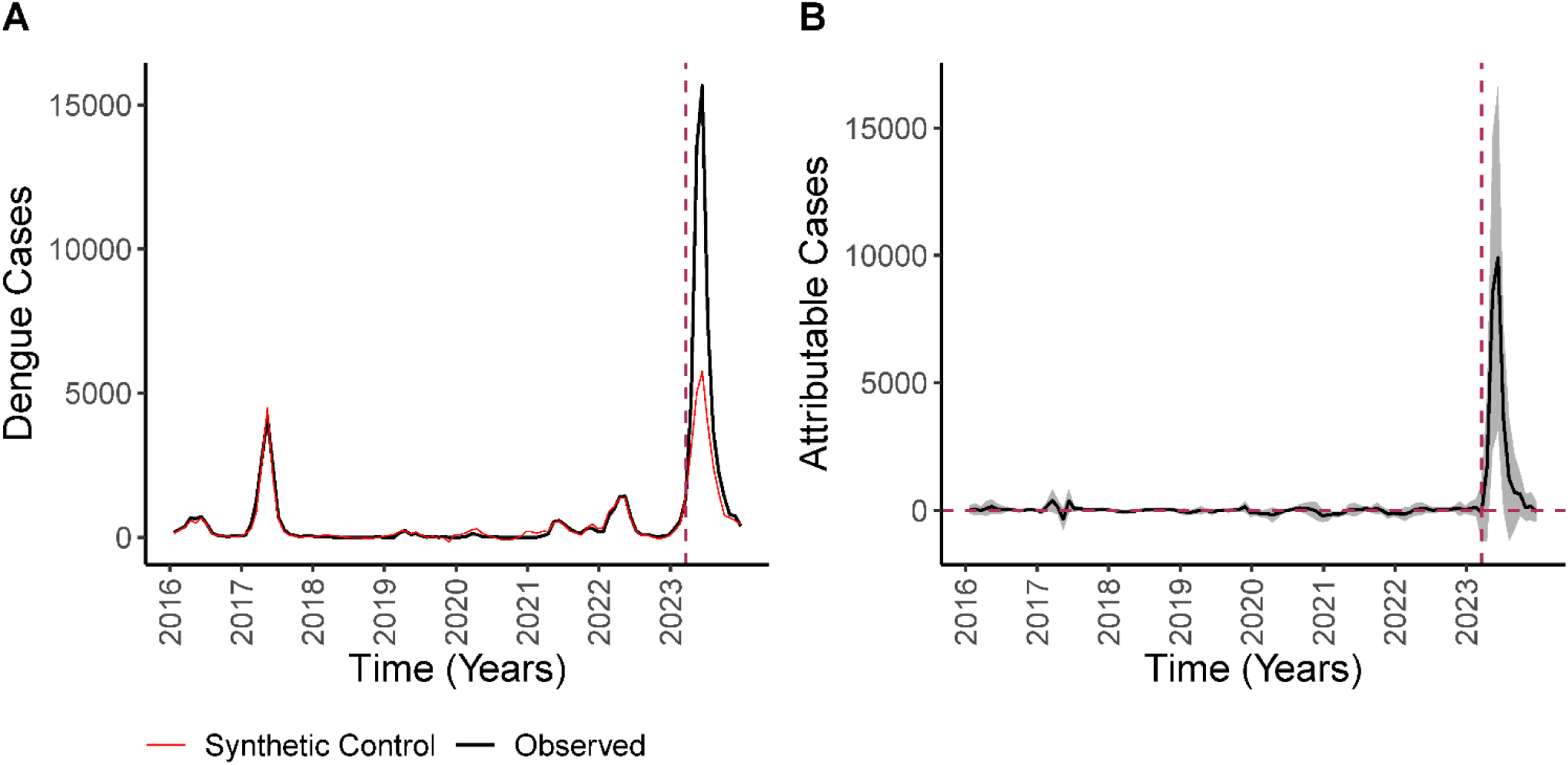
Main results of generalized synthetic control analysis across all districts that experienced extreme precipitation in March 2023. (A) Total observed cases (black) compared to the total cases in the synthetic co trol districts (red) over time (divided into four-week periods) across all districts with extreme precipitation. (B) The difference between observed cases in the districts with extreme precipitation versus their synthetic controls over time, where the period after the dashed line indicates the effect of extreme precipitation during Cyclone Yaku. The grey ribbon corresponds to the 95% confidence interval. The dashed horizontal line indicates no effect and the dashed vertical line indicates when the cyclone occurred, meaning the only the difference in cases to the right of this vertical line is attributable to extreme precipitation during Cyclone Yaku.

Our main estimate of dengue cases attributable to extreme precipitation during Cyclone Yaku was robust to several alternative model specifications. The percentage of attributable cases did not change substantially when temperature was excluded from the model (Figure S9, Figure S10). We estimated a smaller percentage of attributable cases (50%; 95% CI: −25% - 69%) when the control pool included only coastal districts or excluded districts that experienced negative precipitation anomalies during Cyclone Yaku (52%; 95% CI: 11% - 77%) (but in these specifications, our synthetic control model may have been more heavily dependent on districts that were partially affected by the cyclone and coastal El Niño, Figure S11, Figure S10, Figure S12). Estimates were consistent when temperature was transformed (Figure S14) and increased slightly when the time series was extended to include observations prior to 2016, but there was no impact of additionally removing observations from the peak of the COVID-19 pandemic (2020 - 2021; Figure S15, Figure S16). The model fit worsened and the percentage of attributable cases declined when matching was conducted on additional variables (Figure S17). The percentage of attributable cases generally increased when the threshold for non-extreme precipitation decreased and when the upper threshold for extreme precipitation decreased (Figure S18). Estimates were generally robust to the number of control districts to which each district affected by extreme precipitation was matched, the number of latent factors included in the model (Figure S19, Figure S20), and the inclusion of Las Piedras district, which was an outlier in its factor loading values for latent factors 1 and 2, in the matched pool (Figure S21, Figure S22). Consistently, we found that a substantial share of dengue cases was attributable to extreme precipitation.

### 2.3 Effects of extreme precipitation on dengue incidence varied with land use and flood susceptibility

Next, we examined spatial variation in the number of cases that were attributable to extreme precipitation during Cyclone Yaku. First, we noted that the greatest attributable cases occurring in Salitral, Piura (peaking between mid-May and mid-June at 51 attributable cases per thousand and totaling 76.5 attributable cases per thousand people across April 22nd - December 29th) (Figure 3). The northwestern regions of Piura and Lambayeque had several districts with more than 20 cases of dengue per thousand people attributable to extreme precipitation (Figure 3, Figure S1B). Extreme precipitation did not have a large impact on dengue cases in affected districts in San Martin and Cajamarca regions, which are both farther east.

**Figure 3:**
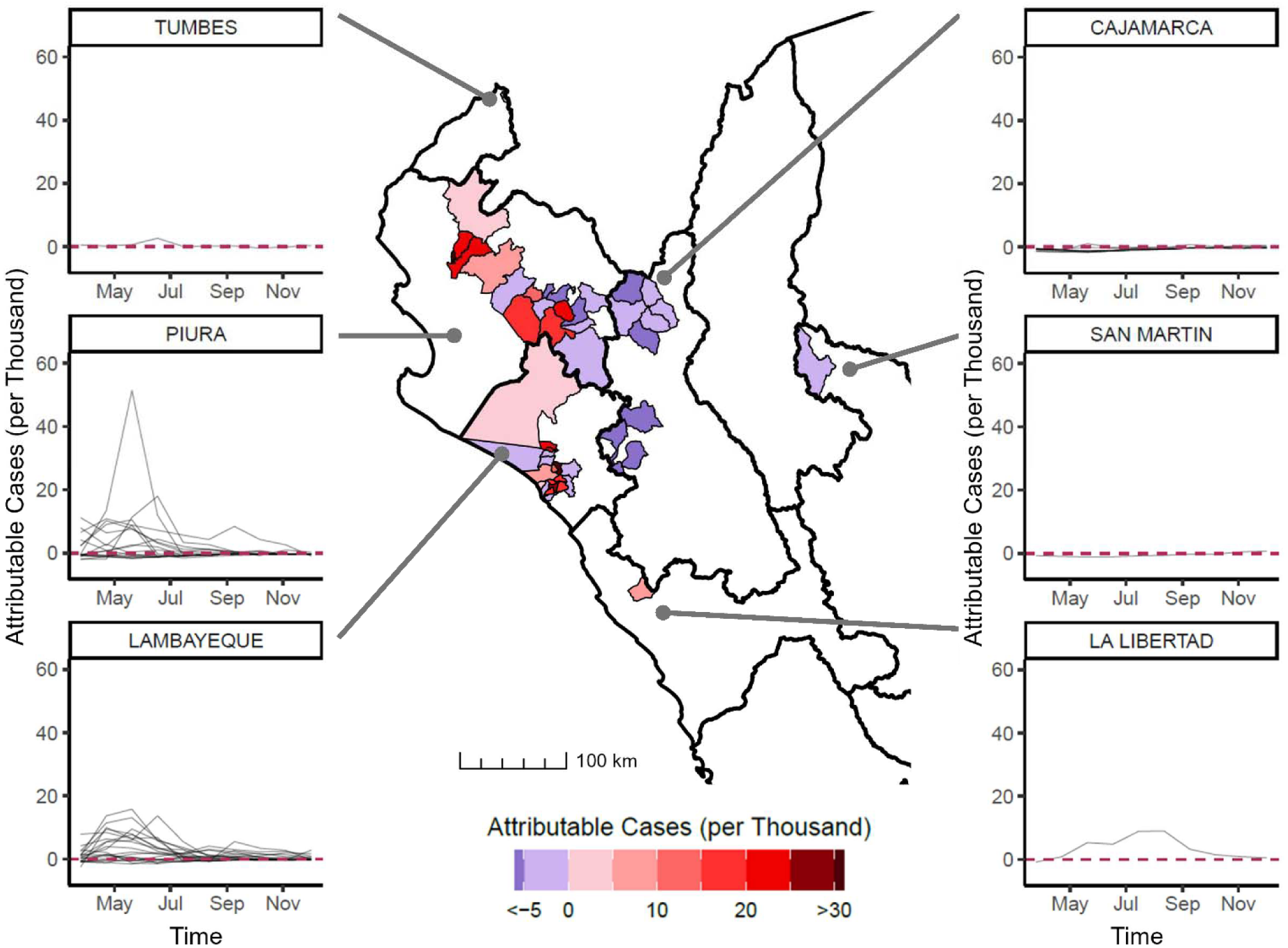
Spatial heterogeneity in the number of dengue cases per thousand people attributable to extreme precipitation. In the center, a map of the number of dengue cases attributable to extreme precipitation per thousand people across districts (calculated from April 22nd - July 14th), zoomed into the region of Peru where extreme precipitation occurred during Cyclone Yaku. Each district affected by extreme precipitation is outlined and shaded according to the estimated number of attributable cases, with darker red indicating a larger positive effect and blue indicating a negative effect. The dark outlines show region borders and lighter outlines show district borders. A scale bar is provided in the bottom left corner of the map. The left and right columns of plots display dengue cases per thousand people attributable to extreme precipitation over time following Cyclone Yaku (in the year 2023) by region, with lines to the map indicating their geographic location. Within the faceted plots, each line corresponds to a different district within the region and the horizontal dashed red lines indicate the baseline of zero dengue cases attributable to extreme precipitation.

We examined a number of potential socio-environmental factors that may modulate the effect of extreme precipitation during Cyclone Yaku on dengue incidence (summarized in Table S5). A single district with extreme precipitation (Lancones, Piura) was excluded from the following analyses because data on all of its vulnerability indices were unavailable. Given evidence of multicollinearity between the vulnerability indices (Figure S24), we summarized multivariate socio-environmental variation among districts using a principal component analysis with rotated components aimed at increasing interpretability.

We tested the relationship between dengue incidence attributable to extreme precipitation during Cyclone Yaku and three rotated principal components (Table S6). Attributable dengue burden was positively associated with the first rotated component (*RC*_1_)(coefficient α_1_ = 5.20, 95% CI: 2.72 - 8.05; *p <* 0.001), which was defined by land use, specifically having more people residing in urban infrastructure and fewer people living in agriculture or pasture land (standardized loadings: 0.94 and −0.92, respectively) (Figure 4, Table S7). Attributable dengue burden was also positively associated with *RC*_3_ (coefficient α_3_ = 6.11, 95% CI: 3.43 - 8.71; *p <* 0.001), which corresponded to with greater flood susceptibility (based on geomorphology and terrain slope) and less household overcrowding (standardized loadings: 0.76 and −0.57, respectively). There was no significant association between dengue incidence attributable to extreme precipitation and *RC*_2_, which was defined by more use of a nonpublic water source and inconsistent water access (coefficient α_2_ = −1.20, 95% CI: −4.25 - 1.76; *p* = 0.232) (Figure 4, Table S7). In other words, the greatest dengue incidence attributable to extreme precipitation occurred in districts with more people residing in land with urban infrastructure, fewer people residing in agricultural or pastoral land, greater flood susceptibility, and less household overcrowding. Emphasizing the importance of warm and wet weather, dengue incidence attributable to extreme precipitation only occurred in districts where mean temperature exceeded 24^◦^C during Cyclone Yaku (Figure 4B).

**Figure 4:**
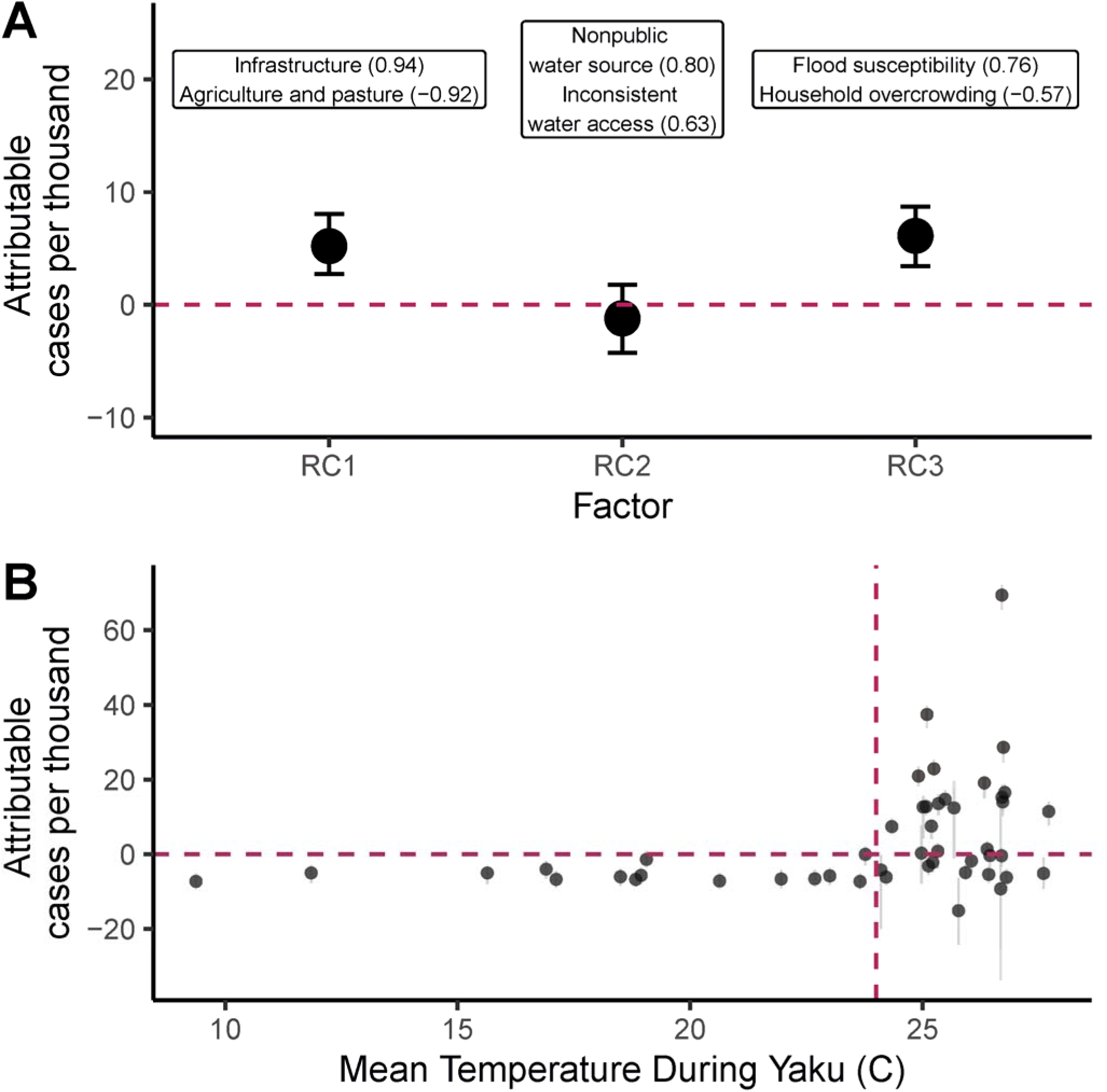
The number of dengue cases attributable to extreme precipitation per thousand people varies across districts with urban infrastructure, flood susceptibility, and temperature. (A) Estimates and 95% confidence intervals for the association between vulnerability indices (rotated components of a principal components analysis of socioenvironmental vulnerability factors, RCs) and dengue cases attributable to extreme precipitation per thousand people at the district level from April 22nd - July 14th. Labels at the top of the plot give the names of the variables most strongly associated with each rotated component (see Table S5 for variable descriptions). The standardized loading is given in parentheses (where | standardized loading | = 1 is the strongest possible association) (see Table S6 for all standardized loadings). (B) The number of dengue cases attributable to extreme precipitation per thousand people (April 22nd - July 14th) is plotted for each district with 95% confidence intervals against the mean temperature in that district during Cyclone Yaku. The red horizontal line indicates no effect of Cyclone Yaku on cases, while the red vertical line indicates 24^◦^C, above which significant attributable cases were observed.

### 2.4 Extreme precipitation coinciding with suitably warm temperatures in March in northwestern Peru is 189% more likely due to historical climate forcing

Climate model simulations of the historical period (1865-2014) suggest that anthropogenic emissions have increased the likelihood of extreme monthly precipitation and warm temperatures suitable for dengue transmission (24^◦^C or greater) during the month of March over northwestern Peru (Figure S25). In accordance with precipitation conditions observed during March 2023 in northwestern Peru, extreme precipitation was defined as mean monthly precipitation exceeding the 84th percentile of observations (Figure S26). Suitably warm temperature was defined as debiased mean monthly temperature exceeding 24^◦^*C*, the threshold above which we observed significantly positive dengue incidence attributable to extreme precipitation (Figure S27, Figure 4B).

Across an ensemble of 203 total realizations of simulated March precipitation values from 7 different climate models, the frequency of extreme March monthly precipitation was 12.24% in the preindustrial period (1865 - 1914) versus 16.00% in the late historical period (1965 - 2014) (Figure 5A). In other words, extreme precipitation was 31% more likely in the late historical period compared to the preindustrial baseline. Based on Kolmogorov-Smirnov tests, the frequency of extreme precipitation events was significantly higher across in the late historical period compared to the preindustrial period (*p <* 0.001). The frequency of suitably warm March monthly temperatures has increased by 109% from 15.70% in the preindustrial period to 32.74% in the late historical period (*p <* 0.001, Figure 5B). Finally, we examined the frequency of concurrent extreme precipitation and suitably warm temperature in March across the region. The frequency of these conditions co-occurring across simulations was 2.57% in the preindustrial period and 7.42% in the late historical period (Figure 5C), constituting a 189% increase (*p <* 0.001).

**Figure 5:**
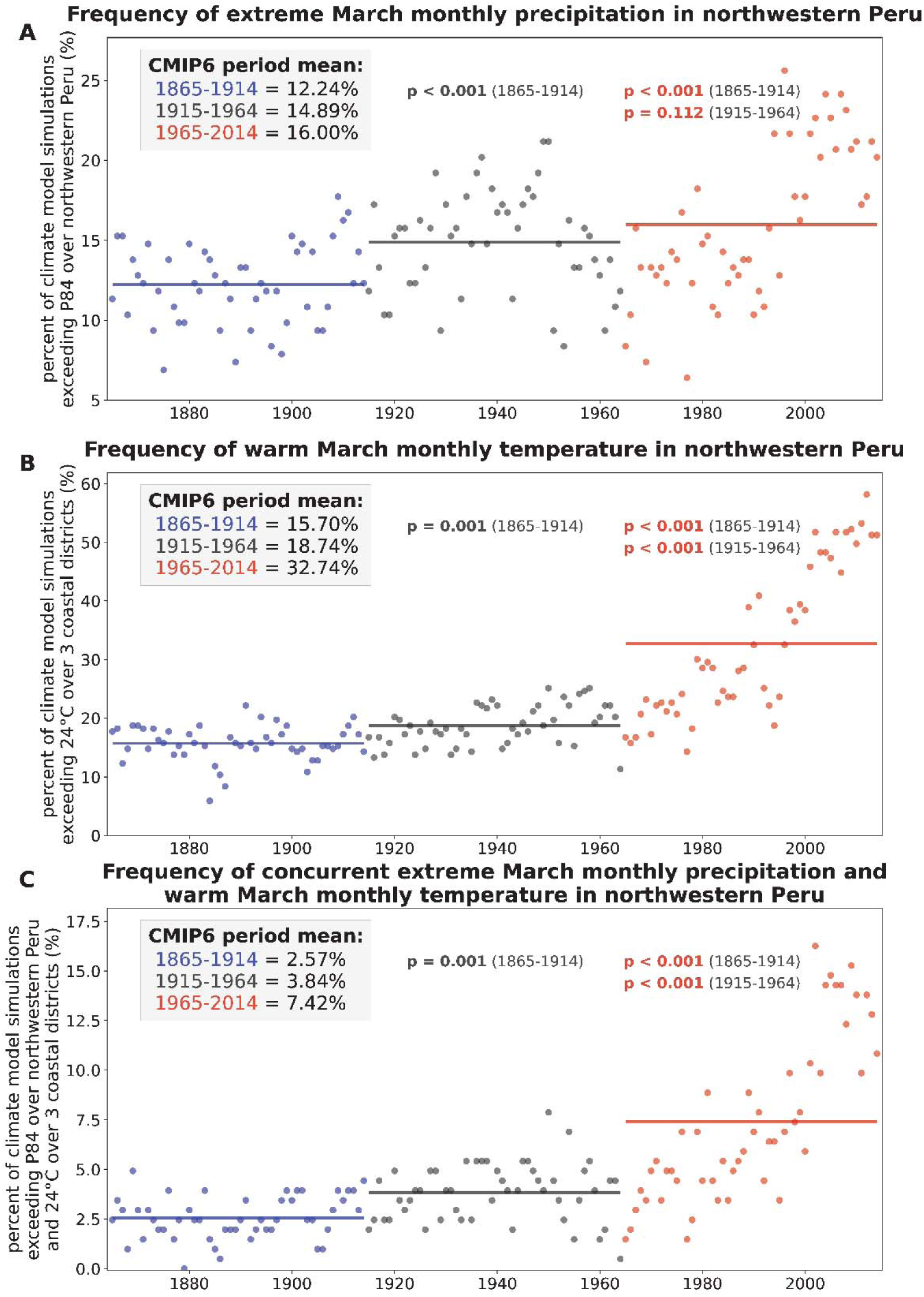
The frequency of extreme precipitation and suitably warm temperature in March in northwestern Peru has increased significantly compared to the preindustrial baseline. Each panel corresponds to a different weather condition: (A) extreme precipitation (B) suitably warm temperature and (C) concurrent extreme precipitation and warm temperature. Annual values show the percentage of 203 climate model simulations exhibiting a given climate condition during each calendar year of the CMIP6 historical forcing experiment for the 1865-2014 period. Based on the quantiles observed in 2023 (see Methods), extreme monthly precipitation is defined to have occurred when the March monthly precipitation averaged over northwestern Peru exceeds the 84th percentile (P84) threshold for the recent climate (calculated from the last 50 years of the CMIP6 historical forcing experiment, or 1965-2014) (Figure S26). Suitably warm temperature is defined to have occurred when March monthly temperature averaged over three coastal regions (Tumbes, Piura, and Lambayeque) exceeds 24^◦^*C*, Figure 4). Solid lines show the mean frequency for the preindustrial (1865-1914, blue), early historical (1915-1964, gray), and late historical (1965-2014, red) periods. We also show p-values for the Kolmogorov-Smirnov (KS) test for the mid- and late-periods calculated relative to the time periods shown in parentheses.

We also compared the likelihood of extreme precipitation and suitably warm temperatures in the early historical period (1915 - 1964) to the preindustrial era. Across these two periods, the frequency of extreme precipitation increased by 22% to 14.89% (*p <* 0.001) and the frequency of suitably warm temperatures increased by 19% to 18.74% (*p <* 0.001). The frequency of these conditions co-occurring across simulations increased by 49% to 3.84% (*p <* 0.001). From the early to late historical period, the frequency of extreme precipitation increased by 7%, a change which was not statistically significant (*p* = 0.11). However, between these periods suitably warm temperatures were 75% more likely and co-occurring warm and wet conditions were 93% more likely, both of which were statistically significant increases (*p <* 0.001). Results were robust to using a percentile-based threshold for suitably warm temperature (subsection S1.17). In sum, the monthly climate conditions observed in northwestern Peru in March of 2023 have become significantly more likely in recent decades due to historical climate forcing.

## 3 Discussion

Anthropogenic climate change is increasing the likelihood and intensity of extreme weather events, which may in turn trigger outbreaks of infectious diseases (Alcayna et al. 2022). Here, we conducted one of the first analyses to connect a climate attribution analysis with a causal assessment of how much a particular extreme weather event contributed to an infectious disease outbreak (Carlson et al. 2024; Ebi and Hess 2020). Over nearly three months, 22,014 (95% CI: 8,665 - 31,712) out of 36,709 dengue cases were attributable to extreme precipitation during Cyclone Yaku in 49 districts of northwestern Peru (Figure 2), corresponding to 60% (95% CI: 24% - 86%) of reported cases and 11.6 (95% CI: 4.6 - 16.7) dengue cases per thousand people across the affected districts. Further, we found that extreme monthly precipitation in March like that associated with Cyclone Yaku in northwestern Peru has become 31% more likely and 189% more likely to co-occur with suitable temperatures for dengue due to climate forcing since the preindustrial era (Figure 5).

Taken together, we found that historical forcing has increased the risk of warm and unusually wet conditions in March in northwestern Peru, which in turn caused the majority of cases during an unprecedented dengue outbreak even after controlling for region-wide increases in dengue. This study builds on existing work linking epidemics to cyclones and other extreme events (Alcayna et al. 2022; Ivers and Ryan 2006). The application of generalized synthetic control methods to estimate a causal relationship is a notable methodological advance given that dengue-focused studies have often been limited in their ability to account for important covariates like immunity, vector control, strain dynamics, and case ascertainment (Laureano-Rosario et al. 2017; Minh An and Rocklöv 2014; Giesen et al. 2020). By showing that the monthly precipitation and temperature conditions that contributed to this outbreak are significantly more likely to occur due to historical climate forcing, we provide some of the first evidence linking mosquito-borne disease burden and climate change (Carlson et al. 2023; Childs et al. 2024). Our climate attribution analysis is consistent with an estimate that anthropogenic forcing increased the likelihood of extreme rainfall across all of Peru in March 2017 by 50% during similar coastal El Niño events (Christidis et al. 2019). This analysis draws attention to the important role of climate change in exacerbating dengue burden in Peru, building on work suggesting that historical warming alone was estimated to have caused a relatively large share of dengue cases in Peru (nearly 40% of recent cases) compared to other countries (Childs et al. 2024). While historical and future warming are expected to increase dengue burden in many parts of the world by increasing temperatures toward the thermal optimum for transmission (Giesen et al. 2020; Messina et al. 2019; Childs et al. 2024), our results highlight the key role of extreme weather in driving epidemics.

Extreme weather events may exacerbate vector-borne disease outbreaks in environments that can support sustained transmission, but timely interventions may preclude transmission. Instances where extreme weather appeared to have varying effects on transmission may be instructive both for forecasting epidemics and for identifying effective preventative measures to apply following extreme events (Beatty et al. 2007; Nosrat et al. 2021). Significant increases in dengue due to extreme precipitation were only observed in districts where mean temperature exceeded 24◦C during Cyclone Yaku, emphasizing that temperature and precipitation conditions combine to determine outbreak risk (Figure 4B). We also found an association between urban infrastructure and higher dengue incidence attributable to extreme precipitation at the district level (Figure 3, Figure 4A, a result consistent with our hypothesis that urban environments have the greatest risk for dengue transmission. We did not find evidence of a significant association between water insecurity and dengue incidence attributable to extreme precipitation, but limited access to clean water has been shown to increase dengue risk in dry settings by promoting water collection and storage in small containers outside of homes (Lowe et al. 2018; Stewart Ibarra et al. 2013; Lowe et al. 2021).

Efforts to increase forest cover, reduce flood risk, and identify and address other environmental risk factors may help reduce dengue burden (Piaggio et al. 2024; Gibb et al. 2023). There is mixed evidence for the role of household overcrowding and other poverty-related factors in determining dengue risk (Mulligan et al. 2015; Borbor-Cordova et al. 2020). Although we do not have data on their implementation within the study districts, protective measures including window screening and covers for water storage containers may help prevent vector contact (Lenhart et al. 2022). Infrastructure improvements focused on land use and urban design may reduce vulnerability to both extreme weather and vector-borne disease (Gibb et al. 2023; Mulligan et al. 2015; Borbor-Cordova et al. 2020).

Our analysis of the effect of extreme precipitation on dengue has several limitations related to baseline differences between the districts with and without extreme precipitation, case reporting, and the lack of an established threshold for extreme precipitation. We addressed potential differences between the groups of districts modeled as cyclone-affected versus control in two ways: first, matching to reduce differences in baseline climate between the two groups of districts and second, weighting the latent factors within the synthetic control model for each district to reconstruct its dengue dynamics prior to the cyclone. Although substantial imbalance remained (Figure S5, Figure S4), matching generally selected for districts with warmer temperatures (creating a bias toward underestimating the effects of extreme precipitation on cases, making our estimate conservative) (Figure S3, Table S2, Figure S5) and improved the fit of the model to actual cases—the key criterion for an acceptable synthetic control (Figure S19). Additional variables (e.g., land use and population density) may be important for dengue dynamics and vary across the districts, but matching on these variables worsened model fit (Figure S17). Although these methods can account for time-varying trends in surveillance and reporting that are shared between groups of districts, our estimate of attributable cases remains sensitive to under-reporting, and all estimates are sensitive to biases in case detection that are not shared by a consistent group of districts. Efforts to expand testing and access to healthcare in under-resourced and crisis settings remain critical, as biases in epidemiological data could lead to uneven and incomplete estimates of the true health costs of extreme weather (Kakkar 2012; Clarke et al. 2024). Finally, we defined extreme precipitation using a relatively conservative threshold of exceeding historical precipitation by more than 8.5 mm/day to ensure a sufficiently large control pool. However, the proportion of cases that were attributable to extreme precipitation was approximately equivalent (68%; 95% CI: 23% - 83%) in a sensitivity analysis using a reduced threshold of 7 mm/day (Figure S18), meaning that our analysis likely underestimates the total number of dengue cases attributable to extreme precipitation by excluding districts with slightly lower precipitation anomalies. Ultimately, our findings were robust to several different model specifications, and weighted sums of the latent factors (i.e., time trends inferred across the matched control districts) were able to reproduce pre-cyclone dynamics in the districts that experienced extreme precipitation during Cyclone Yaku, suggesting that the synthetic control model we constructed produces an appropriate counterfactual (Figure 2).

The climate attribution analyses also had some limitations, most of which are likely to make our results conservative. Although these analyses were designed to analyze the influence of historical climate forcing on precipitation intensity and temperature for districts with the greatest precipitation anomalies during Cyclone Yaku, we averaged our results over a broader region of Northwest Peru through the entire month of March to avoid issues caused by the limited spatial and temporal resolution of available CMIP6 model simulations (Figure S25). In addition, while the CMIP6 archive creates a uniquely large multi-GCM ensemble experiment from which to quantify the influence of historical climate forcing in the context of internal climate variability, the simulations using observed historical climate forcings only extend to 2014. The strong non-stationarity in temperature over this period makes this a conservative choice (since the probabilities are even higher during the most recent period, Figure 5B), as does the fact that greenhouse gas emissions and global warming continued over the subsequent decade after 2014. We also note that the CMIP6 climate models are limited in their ability to realistically simulate extreme precipitation (Donat et al. 2023), causing us to limit our analysis to monthly precipitation during March. Since fine-scale processes can modify extreme event risk (Diffenbaugh et al. 2005), the causal contribution of anthropogenic forcing to precipitation intensity during Cyclone Yaku may be studied more precisely using a high-resolution storyline attribution approach that incorporates the specific atmospheric conditions leading up to this event (e.g., Reed and Wehner (2023); Trok et al. (2024)).

Extreme weather events often have multiple features that may impact human health. For example, Cyclone Yaku also brought heavy wind, mudslides, and widespread flooding to affected areas that likely had additional effects beyond the anomalous precipitation per se (SENAMHI 2023; MapAction). Causal methods may be applied to other instances when extreme weather preceded infectious disease outbreaks to quantify the impacts of several different events and help identify how event characteristics (e.g., wind speed, total rainfall, and event duration) and the extent of resulting damage (e.g., destroyed housing and damaged infrastructure) relate to transmission. Additional analysis across larger regions and multiple events may produce more generalizable results regarding factors that moderate the effects of extreme precipitation on dengue transmission and the relative importance of different potential drivers of dengue risk, which may be measured explicitly and examined using alternative modeling approaches (Gibb et al. 2023; Laureano-Rosario et al. 2017; Nova et al. 2021). Assessing linkages between anthropogenic climate change and health outcomes may guide adaptation efforts while ensuring that the social and environmental costs of fossil fuel emissions are weighed accurately in litigation and climate policy (Stuart-Smith et al. 2021; Burger et al. 2020; Limaye et al. 2020; Ebi et al. 2020; Scovronick et al. 2019). Given the connections that we have demonstrated between historical climate forcing, extreme precipitation, and a large dengue epidemic, mitigating further climate change and building climate-resilient infrastructure may help prevent epidemics and protect human health against further increases in dengue risk.

## 4 Methods

Analyses were conducted in R version 4.2.1 (R Core Team 2022) and Python 3.12.8 (van Rossum 2024). The Stanford University Institutional Review Board determined that this project does not involve human subjects. Code to conduct analyses is available on Github at https://github.com/mjharris95/yaku-dengue.

### 4.1 Climate, case, and vulnerability index data

Mean temperature and total precipitation reported hourly in the ECMWF ERA5-Land Hourly reanalysis dataset at a 0.1^◦^ resolution were extracted at the district level using Google Earth Engine (Copernicus Climate Change Service (C3S) 2017). Given evidence of negative bias in this dataset compared to weather stations, particularly at high elevations, we debiased hourly temperature across pixels following Childs et al. (2024) (see subsection S1.18). We then calculated the daily average temperature and total precipitation across each district by cropping pixels to district boundaries and then taking a population-weighted average across pixels (e.g., weighting by the proportion of the total population living in a given 100m x 100m pixel, estimated for 2020 by WorldPop (Edwards et al. 2021)). In cases where a pixel in the population data was divided by a district boundary, population residing in the pixel was distributed proportionally to the fraction of land area contained in the district for the corresponding pixel.

Weekly case reports (probable and confirmed) by district (administrative division 3) in Peru from 2010 - 2023 were provided by CDC Peru. Probable cases are defined by symptom type and duration and potential exposure (residing in or visiting an area with local transmission of dengue and/or infestation by *Ae. aegypti*, while confirmed cases require laboratory-based testing or (during known outbreaks) an epidemiological link to a known case. The main analysis spanned 2016 through 2023, and a sensitivity analysis included all observations from 2010 onward (subsection S1.8). We focused on the period from 2016 onward to balance including sufficient information from the pre-cyclone period (particularly the size of another large outbreak in 2017) against the potential for the model to become unreliable when fit to too long of a pre-cyclone period (especially given changes to the case definition in 2016) (Xu 2017). Incidence was calculated by dividing reported cases by population size, provided by the Oficina General de Tecnologías del Ministerio de Salud (OGTI, General Office of Information Technology of the Ministry of Health) and estimated annually by Instituto Nacional de Estadística e Informática (INEI, National Center for Statistics and Informatics) based on ten-year national census data. Socio-environmental vulnerability indices for districts with extreme precipitation were accessed through CDC Peru, which compiled data on variables known to be relevant to dengue transmission from other sources (Table S5). The vulnerability indices were: distance to roads, distance to natural bodies of water and rivers, susceptibility to flooding (calculated based on geomorphology and terrain slope, see CENEPRED (2023)), and proportion of residences with: non-public water sources, inconsistent water access, household overcrowding, low-quality construction materials. Land use type was also incorporated as the proportion of the population living in areas classified as infrastructure, agriculture and pasture, or dry forest (from Instituto del Bien Común (IBC) (2022)), the predominant land use types in affected area (Figure S23).

### 4.2 Identifying districts with extreme precipitation

We constrained our analysis to the 561 districts that reported cases of dengue in 2023. Cyclone Yaku, which was first detected by El Servicio Nacional de Meteorología e Hidrología del Perú (the National Service of Meteorology and Hydrology of Peru, SENAMHI) on March 7th, 2023 and dissipated on March 20th, primarily impacted the northwestern coast of Peru (from Tumbes in the north to La Libertad in the south, Figure 1A) (SENAMHI 2023). Given that there was no map of districts most affected by Cyclone Yaku, we focused on districts that received anomalously high precipitation between March 7 - 20, 2023. Anomalies were defined as the difference between mean daily precipitation across this time period in 2023 compared to the historical reference years 1973 - 2022. Averaging over a fifty-year reference period established a recent baseline while also smoothing over recent instances of elevated precipitation (Figure S5). We plotted the distribution of precipitation anomalies across all districts to identify a cutoff value to distinguish districts with extreme precipitation from those with non-extreme precipitation. Specifically, we defined anomalies exceeding 8.5 mm/day as extreme precipitation and anomalies below 7 mm/day as non-extreme precipitation. The cutoff of 8.5 mm/day corresponded to the 90th percentile of the distribution of precipitation anomalies across districts reporting dengue in 2023, a threshold that indicated a relatively large precipitation anomaly during this time in Peru and ensured that a sufficient number of districts could be included in the control pool (Figure S1).

### 4.3 Matching

There are several distinct climatic zones within Peru (from west to east: the dry Pacific coastline, the cool Andes mountains, and the warm and wet Amazon rainforest) (Figure 1B,C), which exhibit substantially different dengue dynamics (Chowell et al. 2011). If the control pool included districts with baseline climate conditions that are substantially different from those of the districts that experienced extreme precipitation, the generalized synthetic control model may overfit to latent trends that are not relevant to the area affected by extreme precipitation during Cyclone Yaku. We therefore used matching to further filter down the set of districts with non-extreme precipitation to a control pool with recent temperature and precipitation trends most similar to the districts with extreme precipitation. We aggregated daily temperature and precipitation to four-week averages to smooth over short-term variation and conducted matching across the 67 observations from the beginning of 2018 until Cyclone Yaku. Using the PanelMatch package we identified the five districts with non-extreme precipitation that were most similar climatically to each district with extreme precipitation based on Mahalanobis distance, a metric that favors units where the relationship between temperature and precipitation is similar to that in the districts with extreme precipitation (Imai et al. 2023; Kim et al. 2022; R Core Team 2022; Mahalanobis 1936). We tested whether the analysis was affected by matching on additional variables, varying the number of control districts to which each district with extreme precipitation was matched, or including all districts with non-extreme precipitation in the control pool without matching (Figure S17, Figure S19).

Districts with non-extreme precipitation that were matched to at least one district with extreme precipitation were included in the control pool used to construct the generalized synthetic control (Figure S2), effectively filtering out districts with non-extreme precipitation that had baseline climate conditions least similar to those across the districts with extreme precipitation during Cyclone Yaku. In a sensitivity analysis, we examined the effects of limiting the potential control pool to districts in coastal regions (Figure S11). To evaluate alignment between the districts with extreme precipitation and the matched control districts, we calculated standardized difference as the difference between each district with extreme precipitation and its matched control districts at a given time point divided by the standard deviation of the corresponding climate variable across all observations.

### 4.4 Generalized synthetic control

We estimated the effect of extreme precipitation during Cyclone Yaku on dengue cases in the affected districts using a generalized synthetic control model (Xu 2017). The dengue incidence (*Y*) in a given spatial unit (*i*) at each four-week time period (*t*) was estimated as:

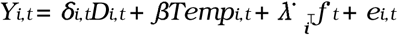

where δ is the effect of extreme precipitation during Cyclone Yaku and *D* is a dummy variable indicating districts with extreme precipitation following the cyclone; Temp is mean population-weighted temperature nine weeks prior to account for biological lags (Nova et al. 2021) with coefficient β; *f*· and λ· are latent factors and factor loadings, respectively, comprising interactive fixed effects (explained in further detail below); and *e* is an error term.

We used interactive fixed effects to control for unobserved time-varying confounders like strain-specific immunity, vector control, case detection, and human movement, which may have different trends and influence across space. A set of n *latent factors* are defined where each latent factor is a time series of constants across the study period. In turn, each spatial unit is assigned a *factor loading*, or vector that weights each latent factor. In other words, the interactive fixed effects term for each spatial unit (*i* = *I*) is a weighted sum across all latent factors k in 1 to n

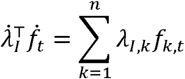

The model was fit to estimate the effects of extreme precipitation in each district over time (δ*_i,t_*) in three steps. First, observations from the control districts alone were used to estimate the coefficient for temperature (β), latent factors (*f*·), and factor loadings (λ·) for the control districts based on the entire time series (including the post-cyclone period). Next, factor loadings were estimated for the units with extreme precipitation such that the mean squared prediction error during the pre-cyclone period is minimized. Because the aim of the generalized synthetic control was to minimize prediction error with respect to cases, factor loadings were determined without any consideration of which control districts were matched to which districts with extreme precipitation based on baseline climate in the prior step. Finally, the effect of extreme precipitation on cases for a given each district *i* in each time period *t* following the cyclone (δ*_it_*) was estimated as the difference between observed incidence and incidence predicted by the synthetic control (*Y*^*_i,t_*):

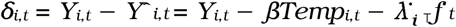

Analyses were conducted using the R package fect: Fixed Effects Counterfactual Estimators (Liu et al. 2022). The number of latent factors to use in the model was selected to minimize mean squared prediction error. For each number of latent factors from zero to five, ten rounds of cross-validation were conducted by withholding 10% of the control units when fitting the model and calculating the mean squared prediction error across the testing units. The number of latent factors that minimized the mean squared prediction error was then selected. Nonparametric confidence intervals for the proportion of cases attributable to extreme precipitation were calculated across 1000 bootstrap runs. We conducted sensitivity analyses to examine the robustness of the model to the following changes in model specifications: excluding temperature as a covariate (Figure S9), limiting the analysis to coastal districts (Figure S11), limiting the control pool to districts with non-negative precipitation anomalies (Figure S12), transforming the temperature covariate based on lab-derived estimates of temperature-dependent transmission rates (Figure S14), including observations prior to 2016 (Figure S15), excluding observations from the peak of the COVID-19 pandemic in 2020 and 2021 (Figure S16), varying the thresholds for extreme and non-extreme precipitation (Figure S18), varying the number of districts with non-extreme precipitation to which each district with extreme precipitation was matched (Figure S19), varying the number of latent factors (Figure S20), and excluding the district Las Piedras which has relatively large factor loadings for factors 1 and 2 (Figure S21, Figure S22).

### 4.5 Examining associations between climate, vulnerability indices, and incidence attributable to extreme precipitation

We examined the relationship between dengue incidence attributable to extreme precipitation and rotated components from a principal components analysis that combines climate covariates and measures of housing quality, water access, land use type (proportion of the population living in land classified as infrastructure, dry forest, or agriculture and pasture, via Instituto del Bien Común (IBC) (2022)), and flood susceptibility, all of which may moderate the effect of extreme precipitation on dengue. We took population-weighted averages of manzana (block)-level vulnerability indices across each district (see Table S5 for a full list and description of socio-environmental vulnerability indices). We also calculated average daily mean temperature and cumulative precipitation during Cyclone Yaku. Given that these socio-environmental vulnerability indices may be strongly correlated with each other (Figure S24), we used the *psych* package in R to load the vulnerability indices across three principal components (Revelle 2007). The principal components were rotated using Varimax rotation, which rotates the principal components so that they more strongly correlate with the variables of interest, improving interpretability (Keith E. Dilbeck 2017). Next, we fit a linear regression of the dengue incidence attributable to extreme precipitation per thousand people in each district (summed from April 22nd - July 14th) against the rotated components (RCs).

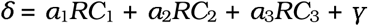

We bootstrapped our estimates of the coefficients (α) and intercept (γ), incorporating uncertainty with respect to dengue incidence attributable to extreme precipitation, by sampling with replacement across districts and their corresponding distributions of estimated dengue incidence attributable to extreme precipitation, interactively fitting parameters and thereby estimating distributions of coefficient and intercept values. Given our strong mechanistic understanding of the relationship between vector biology and temperature (Mordecai et al. 2017, 2019; Lowe et al. 2018), we also examined the relationship between dengue incidence attributable to extreme precipitation and temperature during Cyclone Yaku.

### 4.6 Quantifying the influence of historical climate forcing on the probability of extreme March precipitation in northwestern Peru

To test whether historical climate forcing has significantly increased the likelihood of extreme precipitation coinciding with suitably warm temperatures in northwestern Peru, we analyzed climate model simulations from Phase 6 of the Coupled Model Intercomparison Project (CMIP6) database, which is archived by the Earth System Grid Federation and can be downloaded from their website at https://esgf-node.llnl.gov/search/cmip6/. To incorporate uncertainty arising from both differences in climate model structure and internal climate variability, we downloaded simulations from seven different climate models (ACCESS-ESM1-5, CanESM5, CNRM-CM6-1, IPSL-CM6A-LR, MIROC6, MIROC-ES2L, and NorCPM1) that archived at least 29 realizations in the CMIP6 historical forcing experiment (which spans 1850-2014). Our ensemble thus consists of a total of 203 simulations, each of which provides a unique realization of possible weather conditions that could have occurred during 1850-2014, consistent with the pathway of natural and anthropogenic emissions found in observational data. We then compared the likelihood of extreme precipitation and suitable temperature in March across simulations for three fifty-year periods: 1865 - 1914 (preindustrial baseline), 1915 - 1964 (early historical), and 1965 - 2014 (late historical).

For each of these simulations, we extracted the March monthly mean precipitation data and calculated the area-weighted average over the area of northwestern Peru containing districts affected by extreme precipitation, defined as −3 N to −9 N and −82 E to −76 E (Figure S25). During Cyclone Yaku, March precipitation in regions affected by extreme precipitation exceeded the 84th percentile of observations from 1973 - 2022 according to the population-weighted ERA5 precipitation data (note that this threshold is defined across the distribution of observations *over time*, whereas the 8.5 mm/day threshold was identified based on the distribution of observation *across space* Figure S26). To reflect these conditions, we defined extreme precipitation as a month that exceeds the 84th percentile for monthly mean precipitation during the last 50 years of the historical forcing experiment (1965 - 2014). We calculated the 84th precipitation threshold separately for each climate model to account for biases in the mean and standard deviation of precipitation across different models. To estimate changes in the frequency of extreme March precipitation, we calculated the percentage of simulations with extreme March conditions for each calendar year. We then conducted a Kolmogorov-Smirnov test to examine the likelihood that the observations in each of the two respective later periods came from the same distribution as the observations from the preindustrial baseline. This null hypothesis reflects the assumption that extreme precipitation would have been equally likely in the early and late historical periods compared to the preindustrial baseline without historical anthropogenic emissions.

We repeated this analysis to test whether historical climate forcing has increased the likelihood of suitably warm temperature and the likelihood of concurrent extreme precipitation and warm temperature. In contrast to the precipitation analysis (which uses a percentile-based threshold for extreme events), we defined suitably warm temperatures as months in which the March monthly mean temperature averaged over three coastal districts (Tumbes, Piura, and Lambayeque; Figure S25) exceeds 24^◦^*C*, the temperature threshold above which large dengue outbreaks attributable to extreme precipitation occurred (Figure 3, Figure 4), given the association between absolute temperature and transmission-relevant mosquito biology (Figure S13). To prevent climate model biases from influencing the frequency of months above 24^◦^*C*, we first debiased and downscaled each climate model simulation to match the ERA5 spatial resolution (0.25^◦^ by 0.25^◦^ latitude-longitude grid) using the bias-correction and spatial disaggregation approach (BCSD) (Wood et al. 2004). For each climate model simulation, we extracted the March monthly mean temperature data for 1850-2014 and performed empirical quantile mapping at each grid cell to correct biases between the climate model data and the ERA5 data (bilinearly interpolated to match the climate model grid). Since quantile mapping does not explicitly correct long-term climate trends, we removed the 9-month running mean at each grid cell from the climate model and ERA5 data prior to the bias-correction step, and added these trends back in after quantile mapping (Thrasher et al. 2022). After removing the climate model biases, we spatially disaggregated the climate model data from their native grid to the 0.25^◦^ ERA5 grid by removing the bias-corrected climatology over 1975-2014 from each month, bilinearly interpolating the residuals to the 0.25^◦^ grid, and adding the 0.25^◦^ ERA5 climatology back in to complete the BCSD procedure. We then used this bias-corrected, downscaled climate model data to calculate the area-weighted March monthly mean temperature averaged over three coastal districts (Tumbes, Piura, and Lambayeque) (Figure S27). In a sensitivity analysis, we tested whether results were robust to using a percentile-based threshold for suitably warm temperature (Figure S28, Figure S29).

## 5 Supplemental information

Document S1. Sections S1.1 - S1.18, Figures S1–S29, Tables S1-S7

## Data Availability

All data and code produced are available online at https://github.com/mjharris95/yaku-dengue

## Acknowledgements

This project was made possible by Peru’s Programa Investigando con el CDC (Investigating with the CDC Program). We are grateful to CDC Peru for the opportunities they provide to students through this program. We additionally thank Raisa Paredes and Diego Castro for supporting this collaboration. We thank Kelsey Pano Lyberger for her assistance with obtaining case data for Mexico, Brazil, and Colombia. We thank Marissa Childs for her assistance with extracting climate data from Google Earth Engine. We appreciate the guidance and assistance provided by Yiqing Xu on generalized synthetic control models and the fect package. We also thank Nathan Lo and Andrew MacDonald for their insights on applying synthetic control methods. We would wish to acknowledge computational resources from Google Cloud for Earth Engine and thank Michael Sherman for his assistance applying for additional batch task quota through the Uplift Program. We acknowledge the World Climate Research Programme, which, through its Working Group on Coupled Modelling, coordinated and promoted CMIP6. We thank the climate modeling groups for producing and making available their model output, the Earth System Grid Federation (ESGF) for archiving the data and providing access, and the multiple funding agencies who support CMIP6 and ESGF.

MJH was supported by the Achievement Rewards for College Scientists Scholarship and the National Institutes of Health (R35GM133439). MJH is an investigator at the University of Maryland Institute for Health Computing, which is supported by funding from Montgomery County, Maryland and The University of Maryland Strategic Partnership: MPowering the State, a formal collaboration between the University of Maryland, College Park and the University of Maryland, Baltimore. EAM was supported by the National Institutes of Health (R35GM133439, R01AI168097, R01AI102918), the National Science Foundation (DEB-2011147, with Fogarty International Center), and the Stanford Center for Innovation in Global Health, King Center on Global Development, Woods Institute for the Environment, Sustainability Accelerator, and Institute for Human-Centered Artificial Intelligence. KSM was supported by the Fogarty International Center, National Institute on Aging, and the National Institute of Environmental Health Sciences (NIEHS) under the Global Environmental and Occupational Health program award 5U2RTW010114. AGL was supported by the National Institutes of Health (D43TW007393, with Fogarty International Center). NSD and JTT acknowledge support from Stanford University.

