## Supplemental File for "Extreme precipitation, exacerbated by anthropogenic climate change, drove Peru’s record-breaking 2023 dengue outbreak"

### S1 Supplemental Materials

##### S1.1 Supplemental material: identifying districts with extreme precipitation and matching

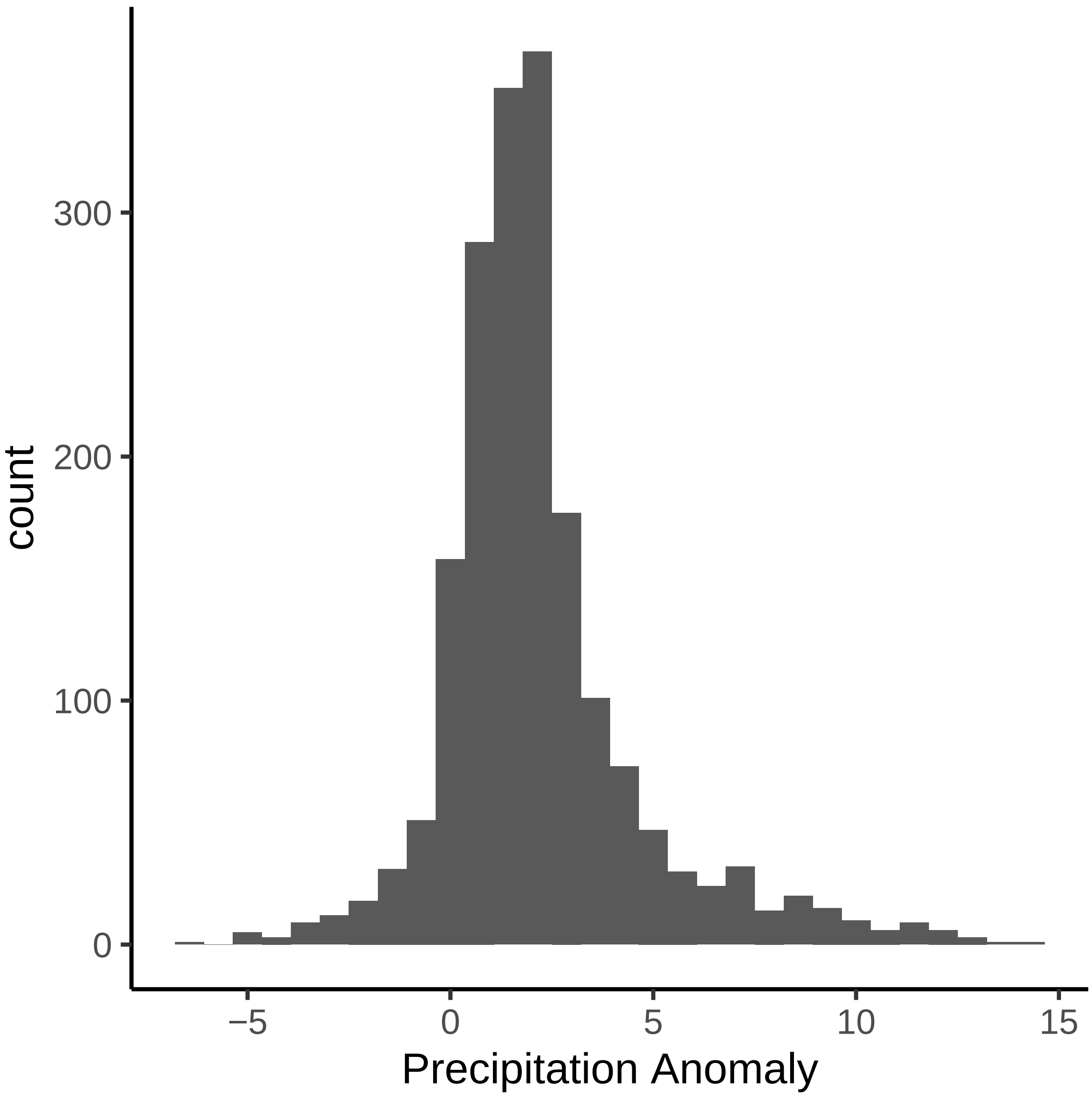

**Figure S1: The distribution of precipitation anomalies (mm/day) during Cyclone Yaku.** Mean daily precipitation from March 7 - 20, 2023 was compared to the historical average between those dates for 1973 - 2022 for all districts in Peru.

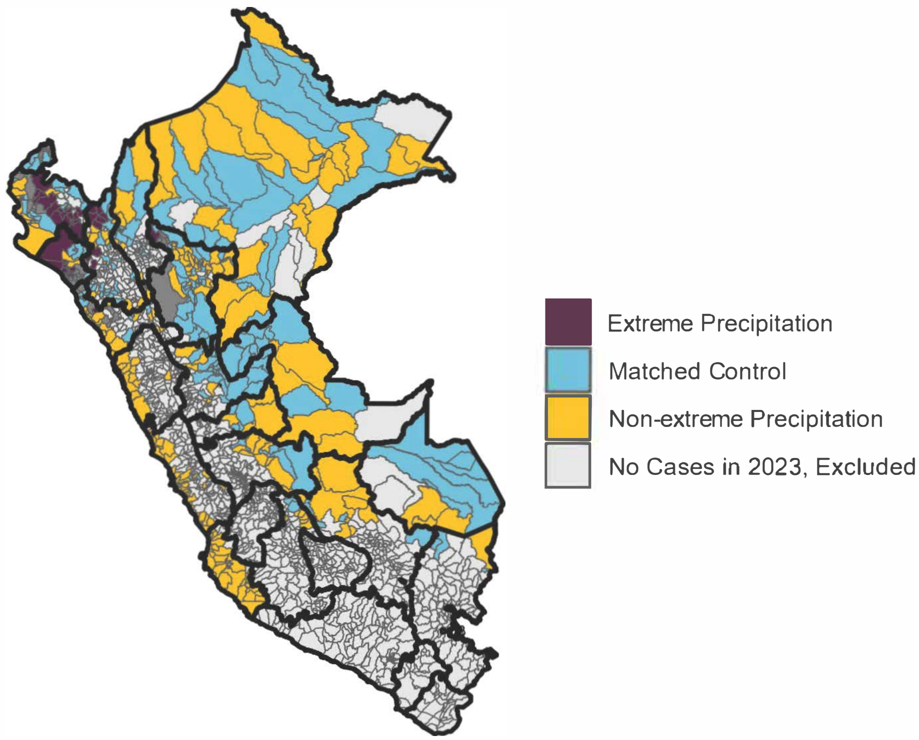

**Figure S2: A map of districts in Peru displaying the extreme precipitation (purple) and matched control (blue) districts.** Eligible units with non-extreme precipitation that were not included in the matched control are shown in yellow. Those excluded from the control set because they did not report cases in 2023 are indicated in light grey while those excluded from the control set because they experienced precipitation anomalies above 7.0 mm/day but below 8.5 mm/day (i.e. buffer districts) are indicated in dark grey. Regional boundaries are indicated with thick black lines.

| Region | Province | District |
|---|---|---|
| CAJAMARCA | CHOTA | LLAMA |
| CAJAMARCA | CUTERVO | QUEROCOTILLO |
| CAJAMARCA | JAEN | SAN JOSE DEL ALTO |
| CAJAMARCA | SAN IGNACIO | SAN IGNACIO |
| CAJAMARCA | SAN IGNACIO | CHIRINOS |
| CAJAMARCA | SAN IGNACIO | LA COIPA |
| CAJAMARCA | SAN IGNACIO | NAMBALLE |
| CAJAMARCA | SAN IGNACIO | TABACONAS |
| LA LIBERTAD | ASCOPE | ASCOPE |
| LAMBAYEQUE | CHICLAYO | CHICLAYO |
| LAMBAYEQUE | CHICLAYO | JOSE LEONARDO ORTIZ |
| LAMBAYEQUE | CHICLAYO | LA VICTORIA |
| LAMBAYEQUE | CHICLAYO | MONSEFU |
| LAMBAYEQUE | CHICLAYO | PICSI |
| LAMBAYEQUE | CHICLAYO | PIMENTEL |
| LAMBAYEQUE | CHICLAYO | REQUE |
| LAMBAYEQUE | CHICLAYO | SANTA ROSA |
| LAMBAYEQUE | CHICLAYO | SAÑA |
| LAMBAYEQUE | CHICLAYO | POMALCA |
| LAMBAYEQUE | CHICLAYO | TUMAN |
| LAMBAYEQUE | FERREÑAFE | FERREÑAFE |
| LAMBAYEQUE | FERREÑAFE | MANUEL ANTONIO MESONES MURO |
| LAMBAYEQUE | FERREÑAFE | PUEBLO NUEVO |
| LAMBAYEQUE | LAMBAYEQUE | LAMBAYEQUE |
| LAMBAYEQUE | LAMBAYEQUE | ILLIMO |
| LAMBAYEQUE | LAMBAYEQUE | MOCHUMI |
| LAMBAYEQUE | LAMBAYEQUE | MORROPE |
| LAMBAYEQUE | LAMBAYEQUE | OLMOS |
| LAMBAYEQUE | LAMBAYEQUE | PACORA |
| LAMBAYEQUE | LAMBAYEQUE | SAN JOSE |
| LAMBAYEQUE | LAMBAYEQUE | TUCUME |
| PIURA | PIURA | TAMBO GRANDE |
| PIURA | HUANCABAMBA | HUANCABAMBA |
| PIURA | HUANCABAMBA | CANCHAQUE |
| PIURA | HUANCABAMBA | SAN MIGUEL DE EL FAIQUE |
| PIURA | MORROPON | CHULUCANAS |
| PIURA | MORROPON | BUENOS AIRES |
| PIURA | MORROPON | LA MATANZA |
| PIURA | MORROPON | MORROPON |
| PIURA | MORROPON | SALITRAL |
| PIURA | MORROPON | SAN JUAN DE BIGOTE |
| PIURA | MORROPON | SANTA CATALINA DE MOSSA |
| PIURA | MORROPON | YAMANGO |
| PIURA | SULLANA | SULLANA |
| PIURA | SULLANA | BELLAVISTA |
| PIURA | SULLANA | LANCONES |
| PIURA | SULLANA | MARCAVELICA |
| PIURA | SULLANA | QUERECOTILLO |
| PIURA | SULLANA | SALITRAL |

**Table S1: Districts with extreme precipitation (continued on next page).** Across each row, the columns indicate the region, province, and district names for each unit.

| Region | Province | District |
|---|---|---|
| SAN MARTIN | RIOJA | ELIAS SOPLIN VARGAS |
| SAN MARTIN | RIOJA | PARDO MIGUEL |
| TUMBES | TUMBES | TUMBES |
| TUMBES | TUMBES | SAN JUAN DE LA VIRGEN |
| TUMBES | ZARUMILLA | ZARUMILLA |
| TUMBES | ZARUMILLA | AGUAS VERDES |
| TUMBES | ZARUMILLA | PAPAYAL |

**Table S1: Districts with extreme precipitation (continued from previous page).**

##### S1.2 Comparing imbalance between districts with and without extreme precipitation before and after matching

Prior to matching, heavier precipitation was associated with relatively hotter temperatures in the districts that experienced extreme precipitation during Cyclone Yaku compared to districts that did not experience extreme precipitation. On average, matching favored control districts where moderate precipitation (e.g., precipitation exceeding 3 mm/day) coincided with hotter temperatures (above 22°C). Matching is therefore important biologically given that dengue risk depends on the co-occurence of suitable temperature and precipitation conditions – particularly heavy rainfall and warm temperatures. The districts with extreme precipitation tended to experience greater fluctuations in temperature compared to the districts without extreme precipitation, although seasonal fluctuations peaked at similar times in both groups (Figure S5B, Table S2). After matching, districts with extreme precipitation during Cyclone Yaku generally had colder temperatures during the dry season compared to the matched control districts but temperatures during the wet season were more similar (meaning that districts with extreme and non-extreme precipitation had similar temperatures during Cyclone Yaku). Districts with extreme precipitation during Cyclone Yaku had less precipitation historically than the control districts, a tendency that remained (and was slightly worsened) after matching (Figure S5A, Table S2). During the cyclone, precipitation in both sets of districts surged but, by definition, deviation from typical conditions was greatest in the districts with extreme precipitation (Figure S5A). Given remaining imbalance, we additionally controlled for temperature within the generalized synthetic control models. A sensitivity analysis demonstrated that matching did improve alignment between predicted and observed incidence ($R^2 = 0.51$ without matching and $R^2 = 0.67$ with matching, see Figure S19).

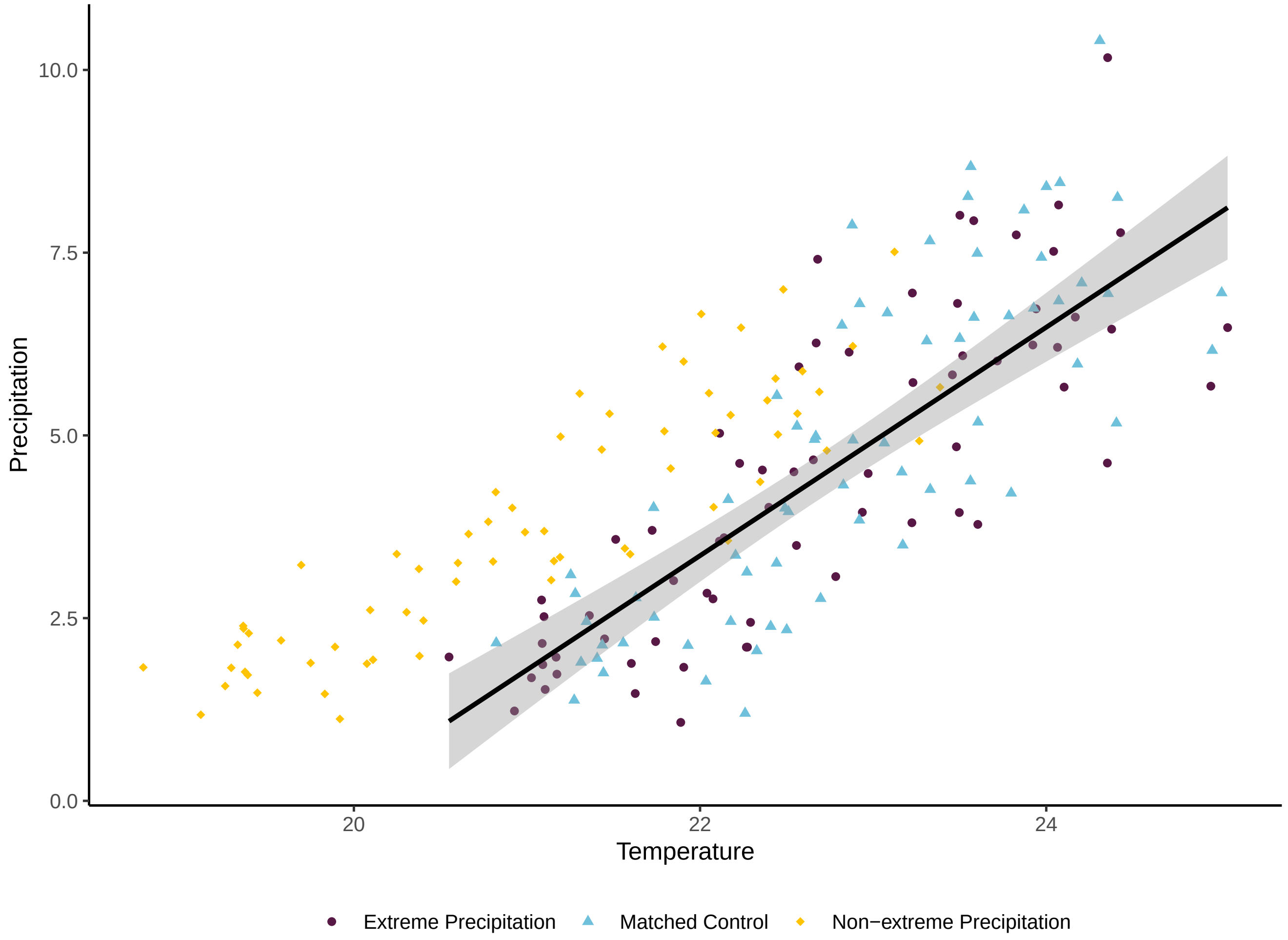

**Figure S3: On average, matching selects for districts where the relationship between temperature and precipitation most closely resembles that in the districts affected by extreme precipitation during Cyclone Yaku**. Each point gives the mean temperature and precipitation at a given pre-cyclone time step across the districts that experienced extreme precipitation (purple, circle), matched control pool of districts with non-extreme precipitation (blue, triangle), and all districts with non-extreme precipitation prior to matching (yellow, diamond). The black trendline indicates the linear relationship between temperature and precipitation in the districts with extreme precipitation and the gray ribbon indicates the 95% confidence interval. For the matched control districts, a weighted average of the climate covariates was calculated with weights corresponding to the number of districts with extreme precipitation to which a given control district was matched. These values are plotted as a time series in Figure S5.

| Group | Mean Temp. (°C) | Sd. Temp. (°C) | Mean Precip. $(\frac{mm}{day})$ | Sd. Precip $(\frac{mm}{day})$ | n |
|---|---|---|---|---|---|
| Extreme precip. | 21.9 | 3.7 | 2.6 | 3.8 | 49 |
| Non-extreme precip. (Full) | 21.5 | 4.8 | 4.3 | 4.3 | 474 |
| Non-extreme precip. (Matched) | 22.6 (23.0) | 4.9 (4.1) | 6.0 (4.8) | 4.6 (4.8) | 123 |

**Table S2:** Covariate balance between the extreme precipitation group (first row) and the non-extreme precipitation group before and after matching (second and third rows respectively). The mean and standard deviation of temperature (°C) and precipitation (mm/day) from 2018 through 2023 are provided. Note, in parentheses, values for the matched control group are weighted by the number of districts with extreme precipitation to which each control district with non-extreme precipitation was matched.

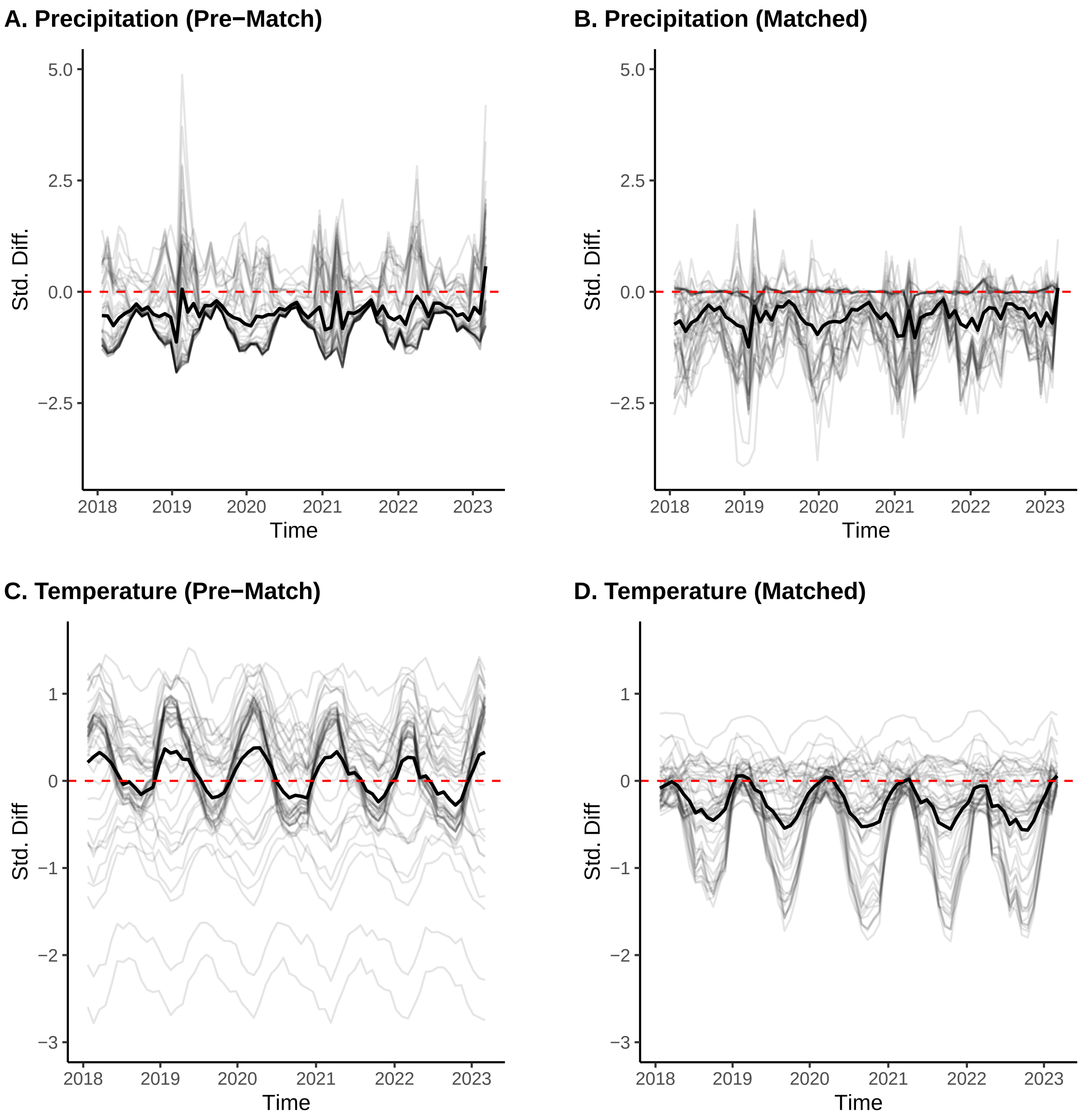

**Figure S4: Covariate balance with respect to precipitation (top) and temperature (bottom) before (left) and after (right) matching**. Within each panel, each line corresponds to a different district with extreme precipitation, comparing its climate covariate values over time to those of either the entire control pool (districts with non-extreme precipitation) prior to matching or the ten control districts to which it was matched. The y-axis corresponds to standardized difference, calculated at each time point as the difference between the value of a given covariate in the districts with extreme precipitation compared to the mean value across its corresponding control districts divided by the standard deviation of each climate covariate over the study period. The horizontal dashed line indicates y = 0, or perfect balance. Observations below the line indicate that the value for a given climate covariate in the control units generally exceeds the value for the units with extreme precipitation (i.e., hotter or wetter conditions in the control units). The thick black line corresponds to the mean standardized difference over time across all districts with extreme precipitation during Cyclone Yaku.

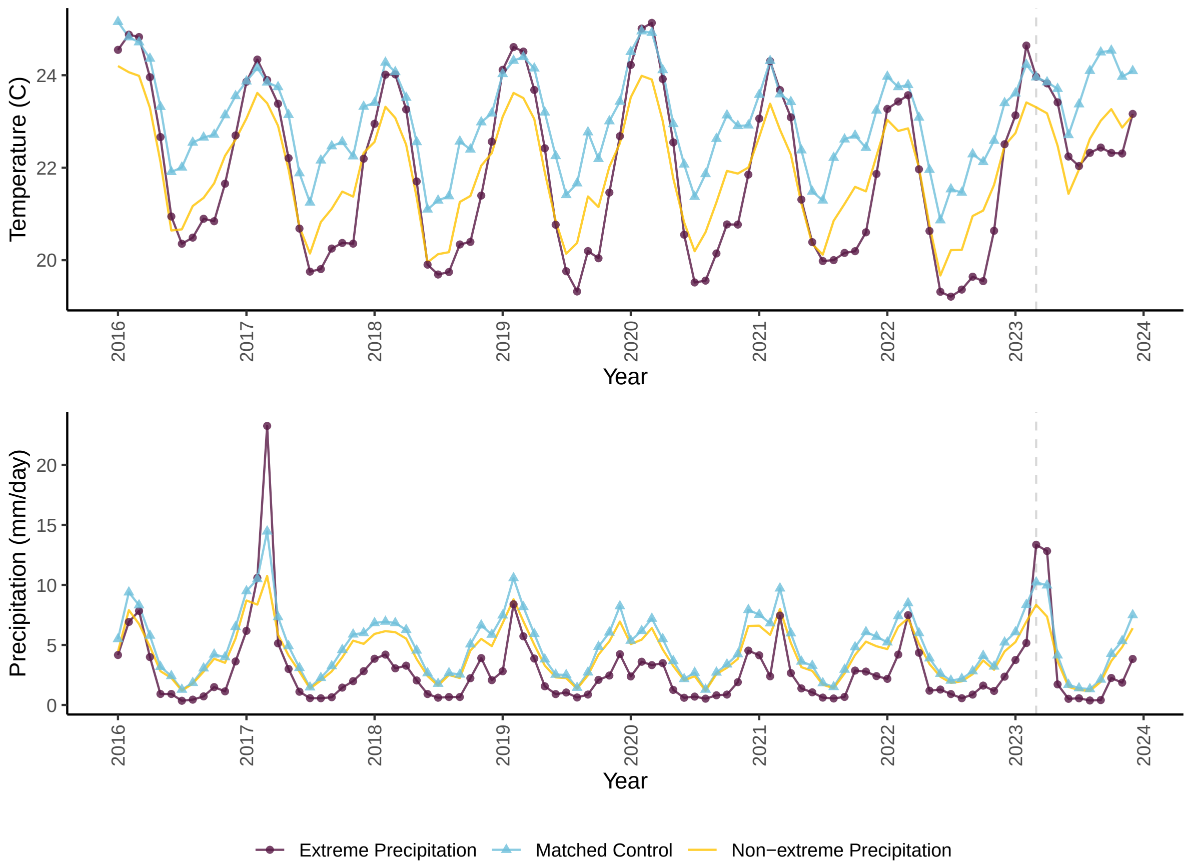

**Figure S5: Climate in the districts with extreme precipitation (purple, circles) compared to the district with non-extreme precipitation before (yellow, solid) and after (blue, triangles) matching.** The top panel shows monthly temperature (°C) and precipitation ($\frac{mm}{day}$) aggregated from 2016 - 2023, with the x-axis indicating time in years. The dotted gray line indicates the month when the cyclone occurred in March 2023. Lines display the mean value of precipitation and temperature across the districts with and without extreme precipitation respectively and a weighted average of the climate covariates across the matched control districts, with weights corresponding to the number of districts with extreme precipitation to which a given control district was matched.

##### S1.3 Supplemental material: generalized synthetic control analysis

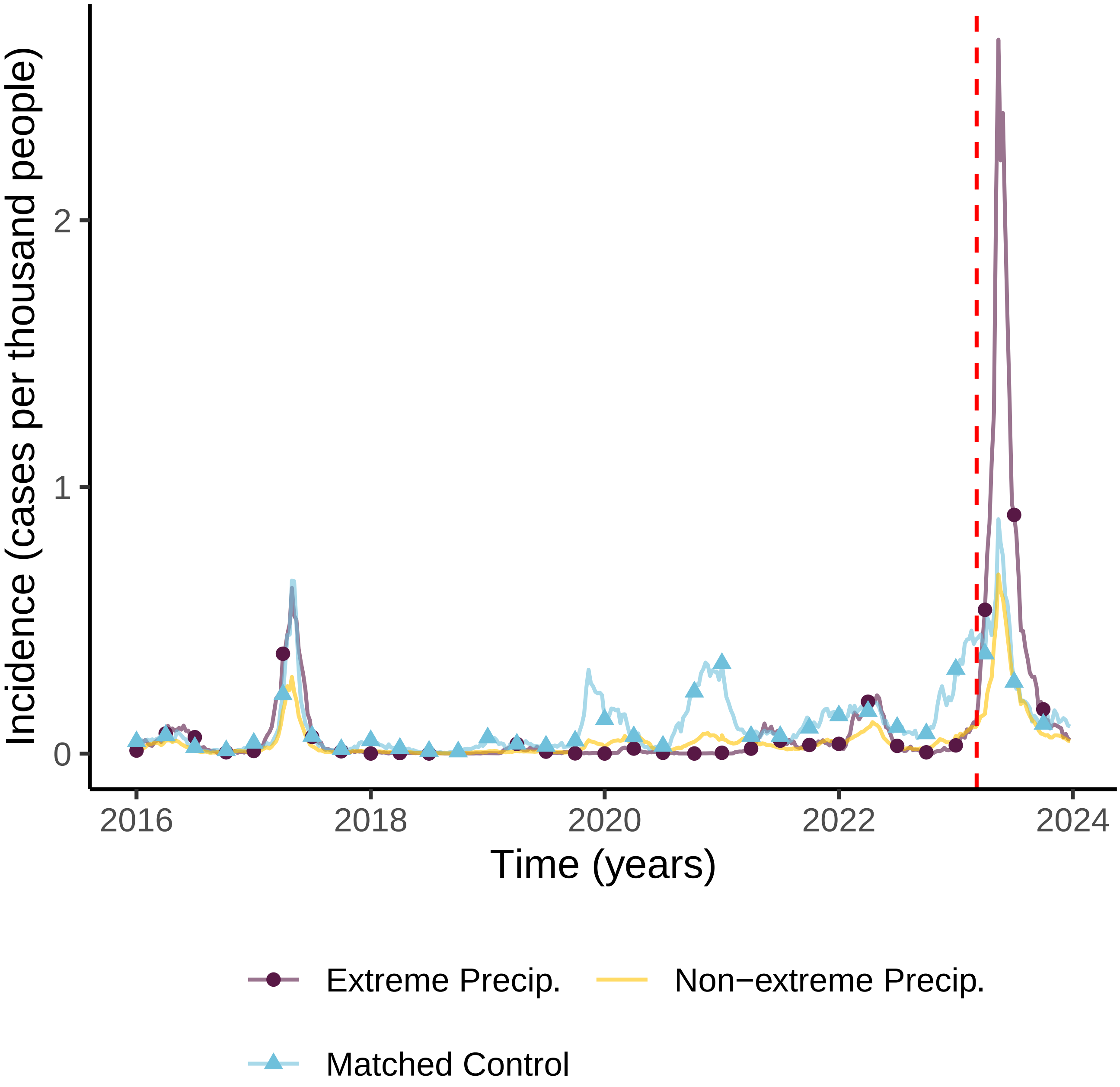

**Figure S6: Dengue incidence across the districts that experienced extreme precipitation during Cyclone Yaku was substantially elevated following the cyclone.** We compare weekly reported dengue cases per thousand people over time in the districts with extreme precipitation (purple, circles) versus the districts with non-extreme precipitation before (yellow, solid) and after (blue, triangles) matching. The dashed red line indicates March 7, 2023, when Cyclone Yaku began.

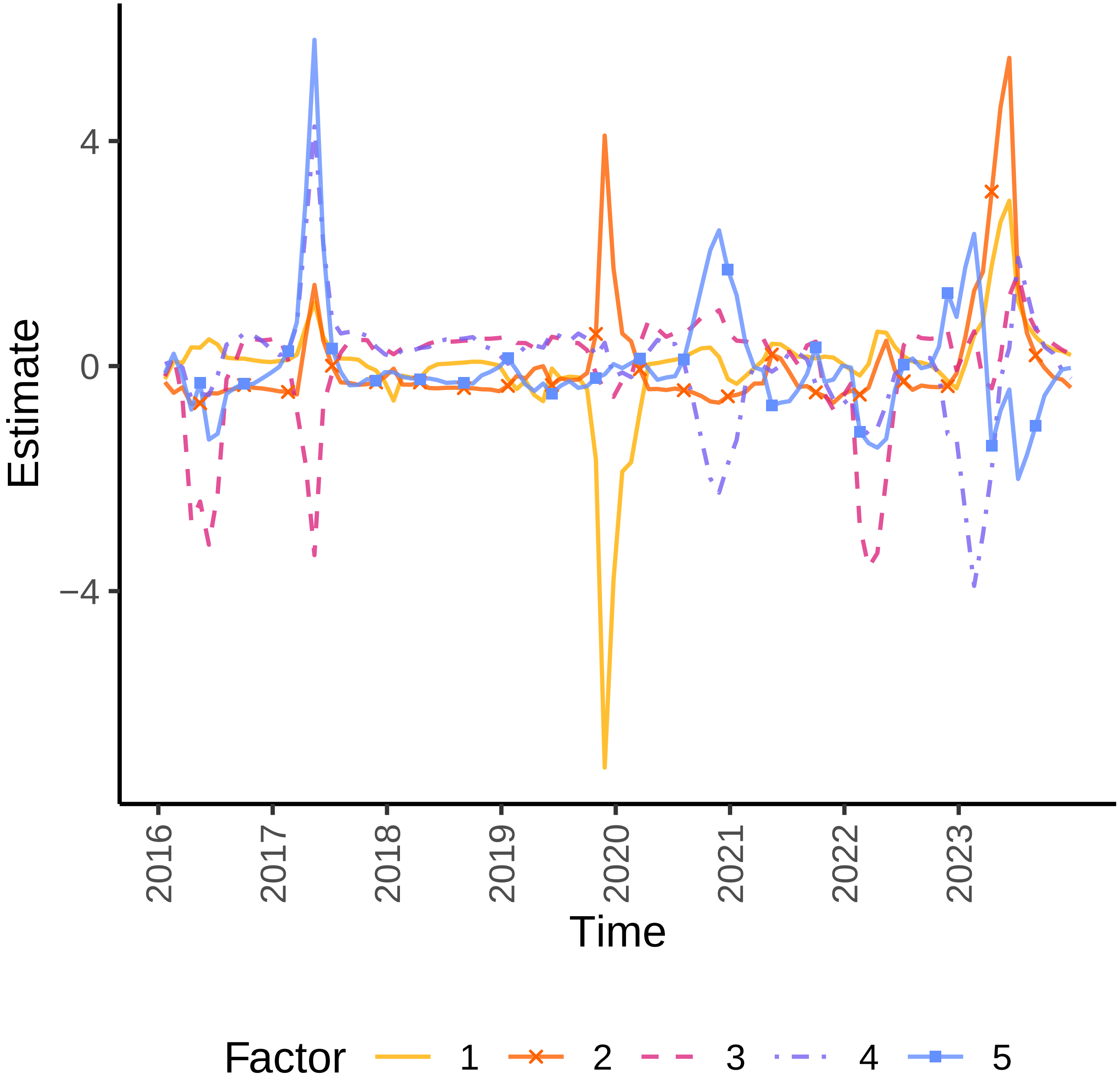

**Figure S7: Latent factors estimated from trends in the control districts, accounting for temperature.** The value of each latent factor (calculated across four-week periods) is plotted over time (x-axis) and each of the five latent factors is indicated with a different color and line type.

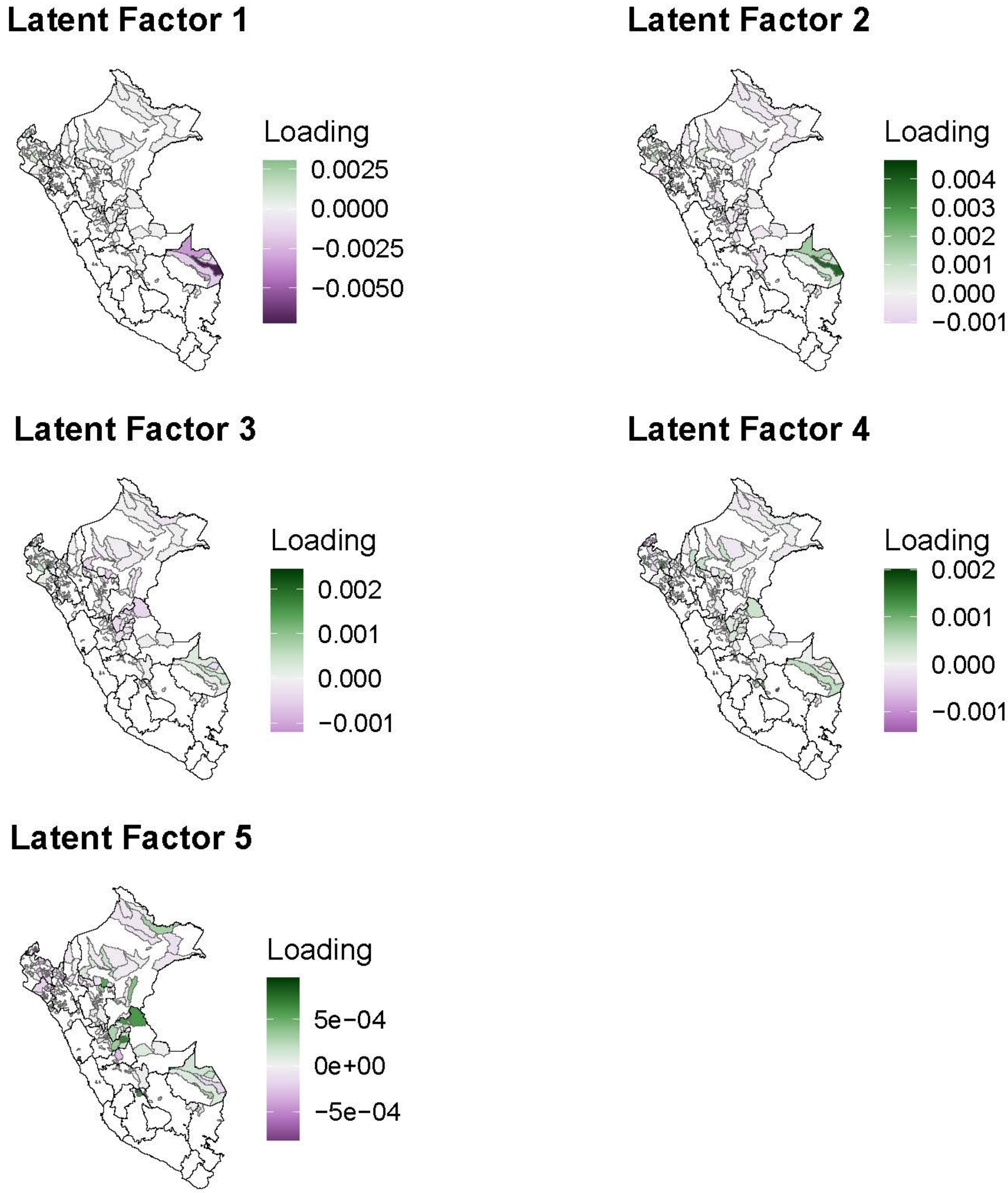

**Figure S8: Maps of factor loadings across districts included in the analysis.** Panels are maps of Peru with dark grey lines indicating regions and black lines indicating districts included in the analysis. Each panel displays the factor loadings for a different latent factor, numbered according to Figure S7, Darker shades of green indicate greater positive values whereas darker shades of purple indicate greater negative values.

| Covariate | Coefficient | p-value |
|---|---|---|
| Temperature (°C) | $3.55 * 10^-5$ ($7.71 * 10^-6$ - $6.34 * 10^-5$) | 0.012 |

**Table S3: Coefficient ($\beta$) estimated for mean temperature.** We additionally provide a bootstrapped 95% confidence interval and p-value for the estimate.

| Dates | Percent Attributable Cases | Number Attributable Cases | Obs. Cases | p-value |
|---|---|---|---|---|
| **Pre-cyclone** | | | | |
| Dec 31 - Jan 27 | 26 (-175, 93) | 81 (-556 - 293) | 317 | 0.424 |
| Jan 28 - Feb 24 | 19 (-137, 72) | 113 (-796 - 419) | 580 | 0.461 |
| **Cyclone** | | | | |
| Feb 25 - Mar 24 | -7 (-135, 74) | -90 (-1804 - 988) | 1336 | 0.635 |
| **Post-cyclone** | | | | |
| Mar 25 - Apr 21 | 36 (-36, 85) | 1744 (-1760 - 4113) | 4846 | 0.131 |
| Apr 22 - May 19 | 63 (13, 88) | 8535 (1785 - 11981) | 13554 | 0.007 |
| May 20 - Jun 16 | 63 (28, 91) | 9918 (4434 - 14191) | 15673 | 0.003 |
| Jun 17 - Jul 14 | 48 (6, 94) | 3561 (444 - 7041) | 7482 | 0.017 |
| Jul 15 - Aug 11 | 33 (-42, 104) | 1233 (-1539 - 3854) | 3703 | 0.108 |
| Aug 12 - Sep 08 | 32 (-33, 100) | 707 (-729 - 2246) | 2237 | 0.113 |
| Sep 09 - Oct 06 | 46 (-2, 113) | 635 (-26 - 1579) | 1395 | 0.031 |
| Oct 07 - Nov 03 | 14 (-51, 76) | 113 (-400 - 599) | 787 | 0.256 |
| Nov 04 - Dec 01 | 21 (-56, 96) | 155 (-411 - 709) | 735 | 0.209 |
| Dec 02 - Dec 29 | -15 (-137, 77) | -57 (-530 - 299) | 387 | 0.587 |

**Table S4: Estimated effects of extreme precipitation on dengue over time impacted districts**. In order, the columns indicate: the start and end date of the time period (in 2023) across which the effects of extreme precipitation were estimated (note that the first time period begins on December 31, 2022) with two pre-cyclone periods and the period when the cyclone occurred included for comparison; the percent of total cases in the affected districts attributable to extreme precipitation with the 95% confidence interval in parentheses; the estimated number of additional cases of dengue caused by extreme precipitation across the affected districts with the 95% confidence interval in parentheses (negative values indicate decreases in cases due to extreme precipitation); the total observed cases across the districts with extreme precipitation during the corresponding period; and the p-value of the estimated effect of extreme precipitation.

#### S1.4 Sensitivity to exclusion of temperature covariate from model

We repeated the main analysis for districts in Peru without including temperature as a covariate in the generalized synthetic control model. Cases were significantly increased by extreme precipitation from April 22nd - July 14th, as was the case for the main model. Overall, we estimate 22,447 (95% CI: 9,256 – 31,949) dengue cases were attributable to extreme precipitation, or 61% (95% CI: 25% - 87%) of all cases reported across the districts with extreme precipitation during this time period (Figure S9, Figure S10B). Unlike the main model, this model does not generally predict continued fluctuations in case counts during periods of low incidence (Figure S9). The $R^2$ of this model was equivalent to that of the main model (Figure S10A).

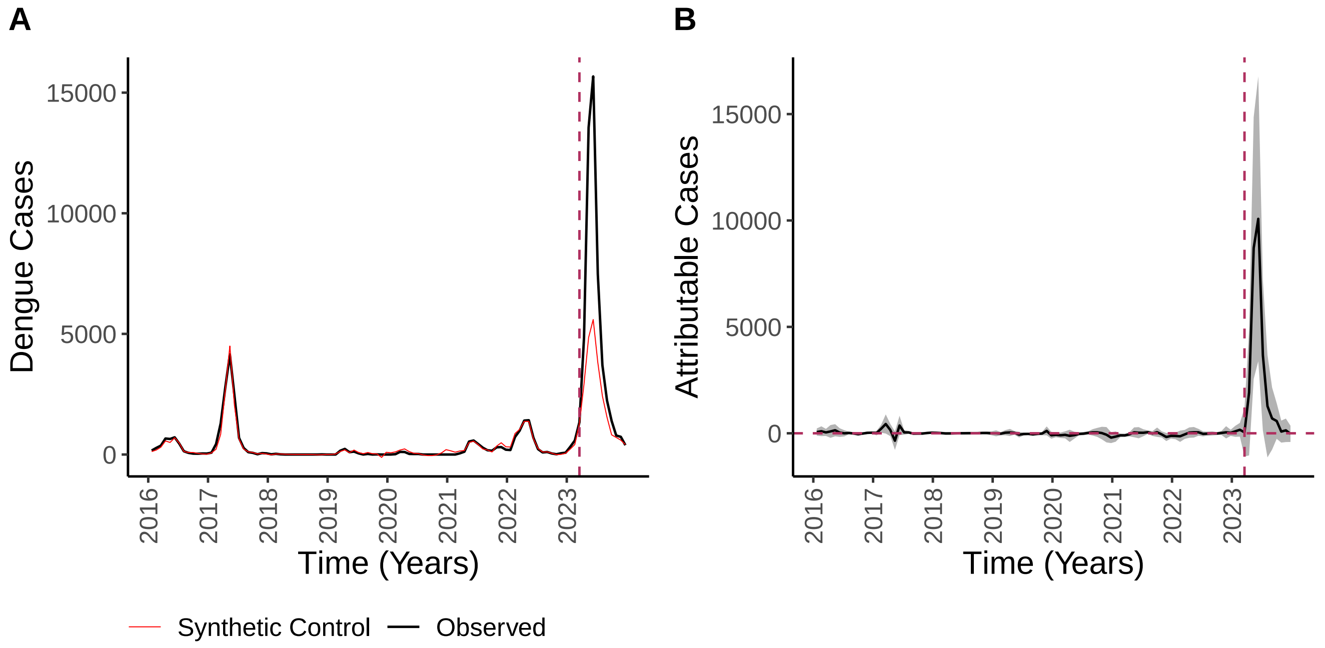

**Figure S9: Results of generalized synthetic control analysis without accounting for temperature.** (A) Shows the total observed cases (black) across all districts with extreme precipitation over time compared to the total cases in the synthetic control (red). (B) Shows the effect of extreme precipitation over time, estimated as the difference between observed cases and synthetic control cases, with the grey ribbon corresponding to the 95% confidence interval. The dashed horizontal line indicates no effect and the dashed vertical line indicates when the cyclone occurred.

#### S1.5 Sensitivity to exclusion of non-coastal districts from control pool

This analysis was limited to districts within coastal departments (Tumbes, Piura, Lambayeque, Cajamarca, La Libertad, Ancash, Callao, Lima, Ica, Arequipa, Moquegua, and Tacna) (Figure S2). The pool of districts with extreme precipitation was reduced to 48 districts because a district in San Martin that was identified as experiencing extreme precipitation in the main text was dropped from this analysis. There were 241 coastal districts without extreme precipitation and 63 matched control districts in this analysis.

The effect of extreme precipitation on cases was never significant ($p < 0.05$) and confidence intervals encompassed negative values across the post-cyclone period. Confidence intervals for this model were generally considerably broader compared to the confidence interval for the model from the main analysis. Across the period that cases were elevated because of extreme precipitation in the main analysis (April 22nd - July 14th), we estimate 18,258 (95% CI: -9,201 - 25,508) cases were attributable to extreme precipitation, constituting 50% (95% CI: -25% - 69%) of all cases. This is likely a underestimate of the effects of extreme precipitation during the period because many coastal districts did experience some impacts of Cyclone Yaku and the coastal El Niño, including heavy precipitation, worsening the control. Further, there may have also been considerable importation of cases from the districts with extreme precipitation into neighboring coastal districts. This model had a substantially lower $R^2$ compared to the model fit to districts across all of Peru ($R^2 = 0.28$) (Figure S10A).

Excluding temperature as a covariate from the model focused on coastal districts had minimal impact on the results (Figure S10B). There was no effect of additionally excluding climate covariates on $R^2$ ($R^2 = 0.28$) (Figure S10A). Again, the effect of extreme precipitation on cases was not significant across the post-cyclone period. Between April 22nd and July 14th, a total of 18,168 (95% CI: -8,801 - 24,072) cases were attributable to extreme precipitation (49% of cases; 95% CI: -24% - 66%) (Figure S10B).

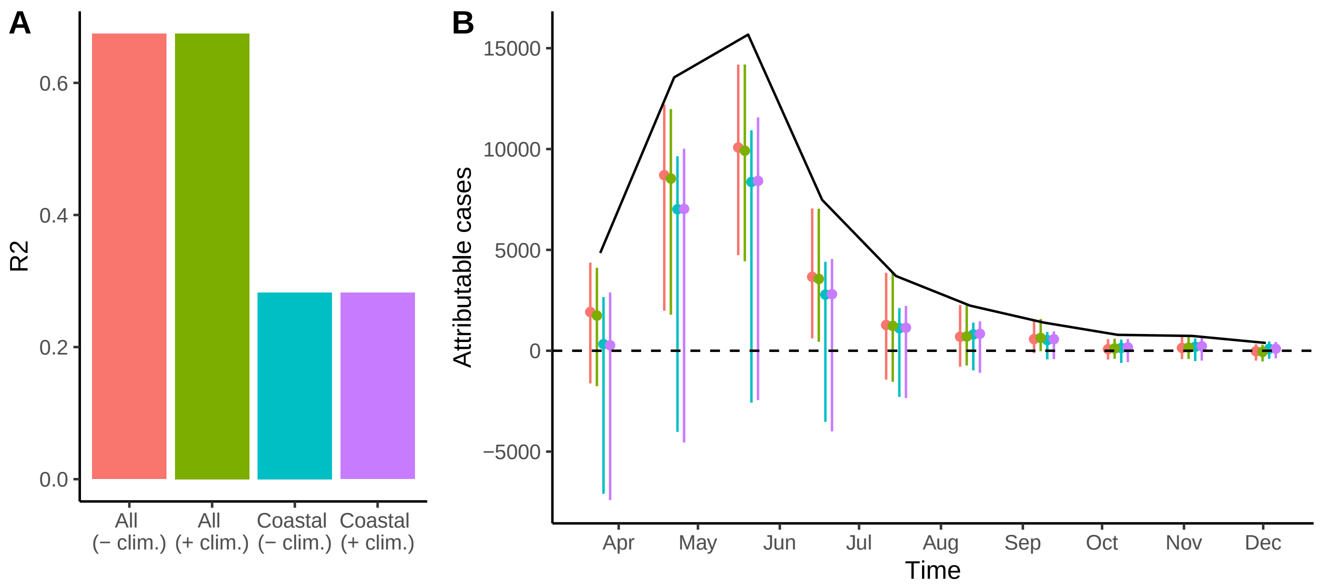

**Figure S10: Comparing synthetic control model fit and results depending on whether coastal districts were included in the control pool and whether climate covariates were included in the model.** In both panels, we compare models with: all districts with non-extreme precipitation in the control pool and temperature excluded from the model (red), all districts with non-extreme precipitation in the control pool and and temperature included in the model (green, main model), only coastal districts with non-extreme precipitation in the control pool and temperature excluded from the model (blue), and only coastal districts with non-extreme precipitation in the control pool and temperature included in the model (purple). (A) Shows the $R^2$ of the generalized synthetic control model depending on model specifications. (B) Shows the estimated effect of extreme precipitation on cases over time (beginning in late March 2023), with the 95% confidence interval. The black line indicates actual cases reported over time in all districts with extreme precipitation, including one that is not coastal. The dashed vertical line indicates no effect of extreme precipitation on cases.

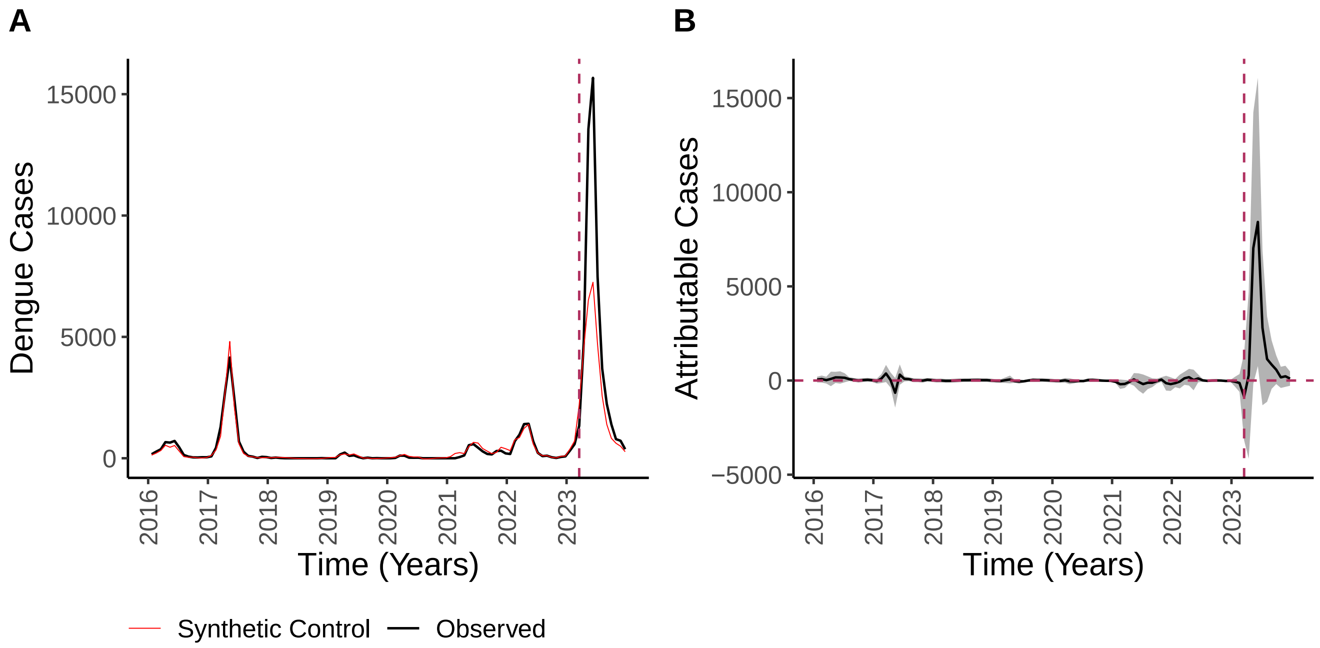

**Figure S11: Results of generalized synthetic control analysis across coastal districts.** (A) Shows the total observed cases (black) across all districts with extreme precipitation over time compared to the total cases in the synthetic control (red). (B) Shows the effect of extreme precipitation over time, estimated as the difference between observed cases and synthetic control cases, with the grey ribbon corresponding to the 95% confidence interval. The dashed horizontal line indicates no effect and the dashed vertical line indicates when the cyclone occurred.

#### S1.6 Sensitivity to exclusion of control districts with negative precipitation anomalies

Given that unusual dryness may also impact dengue transmission dynamics, we tested a model where districts with negative precipitation anomalies (precipitation below the historical daily mean) during Cyclone Yaku were excluded from the pool of districts with non-extreme precipitation eligible to be included in the control pool (Figure 1A). There were a total of 377 districts with non-extreme precipitation and non-negative precipitation anomalies, of which 107 were matched to the cyclone-affected districts.

Cases were significantly increased by extreme precipitation from April 22nd - June 16th, a slightly abbreviated period compared to the main analysis. From April 22nd - July 14th, 19,074 (95% CI: 3,868 - 28,187) cases were attributable to extreme precipitation, constituting 52% (95% CI: 11% - 77%) of cases. The model predictive accuracy was comparable to that of the main analysis ($R^2 = 0.68$).

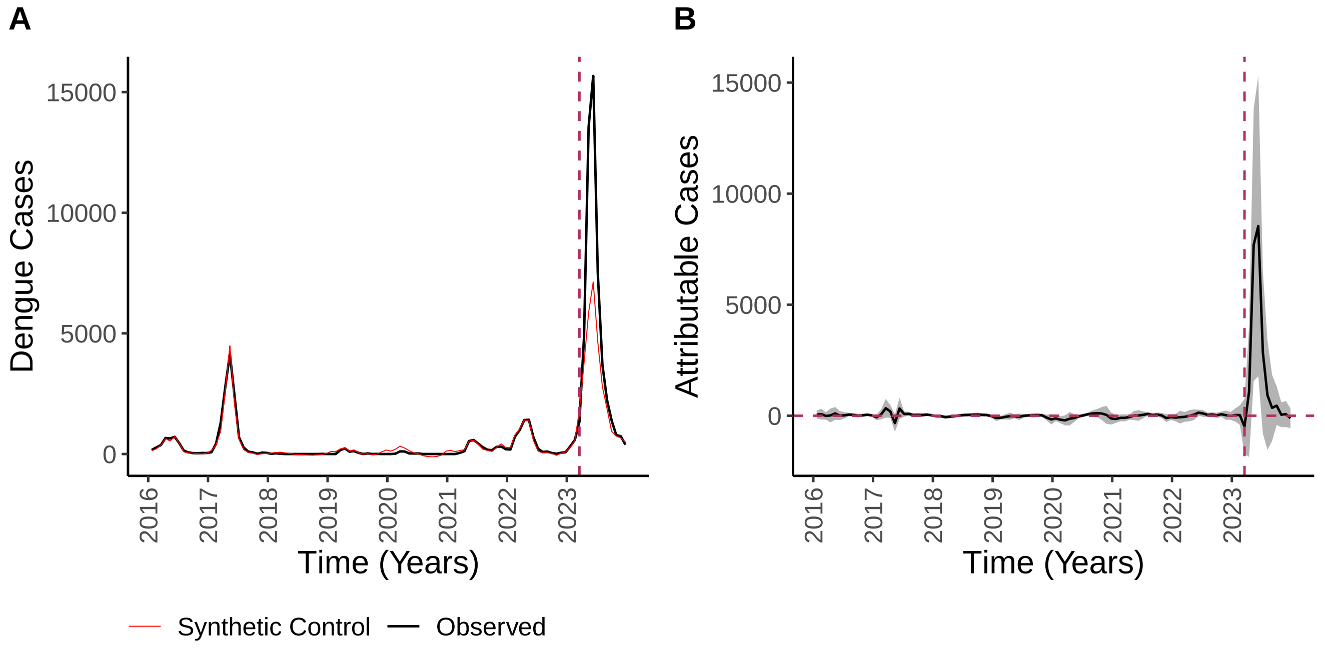

**Figure S12: Results of generalized synthetic control analysis excluding districts with negative precipitation anomalies from the potential control pool.** (A) Shows the total observed cases (black) across all districts with extreme precipitation over time compared to the total cases in the synthetic control (red). (B) Shows the effect of extreme precipitation over time, estimated as the difference between observed cases and synthetic control cases, with the grey ribbon corresponding to the 95% confidence interval. The dashed horizontal line indicates no effect and the dashed vertical line indicates when the cyclone occurred.

#### S1.7 Sensitivity to use of temperature-dependent $R_0$ as climate covariate in model

Prior work has used laboratory measurements of transmission-relevant traits across a thermal gradient to identify a nonlinear relationship between temperature and relative transmission intensity for dengue (Mordecai et al. 2017). We repeated the main analysis for districts in Peru using temperature-dependent relative $R_0$ instead of mean temperature as a covariate in the generalized synthetic control model.

Cases were significantly increased by extreme precipitation from April 22nd - July 14th, as in the main analysis Figure S14. We estimate that 21,632 (95% CI: 8,475 - 32,116) or 59% (95% CI: 23% - 87%) of cases were attributable to extreme precipitation during that time period. These results were similar to those from the main analysis, likely because mean temperature in the study region was generally between 20 and $26°C$, a range where the relationship between temperature and relative $R_0$ is expected to be approximately linear.

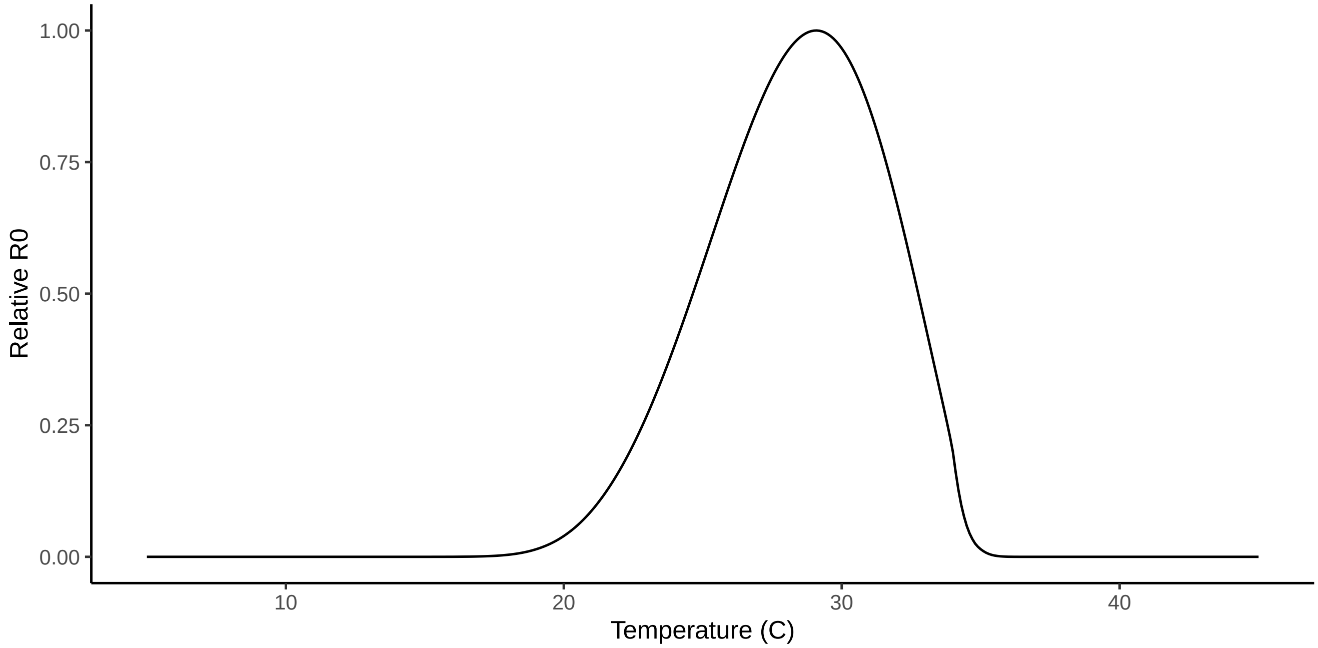

**Figure S13: The relationship between temperature (°C) and relative $R_0$.** Relative $R_0$ is a unitless measure of relative transmission intensity. This relationship was derived by Mordecai et al. (2017) based on laboratory experiments.

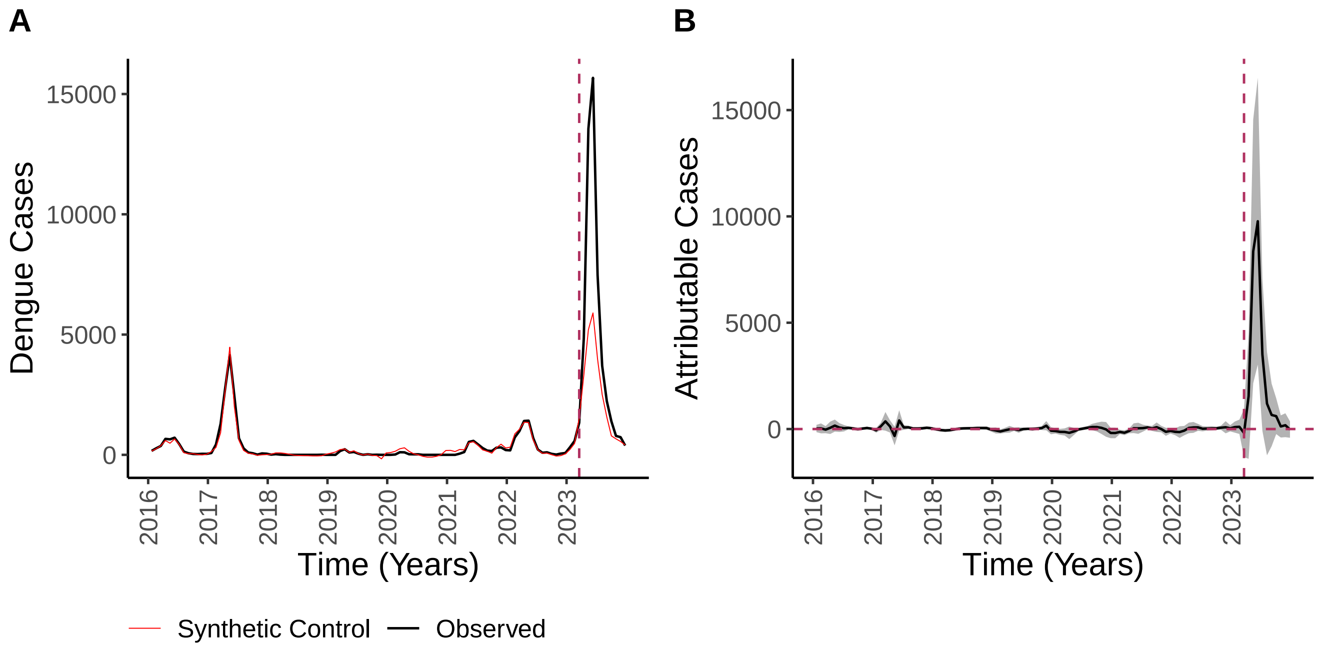

**Figure S14: Results of generalized synthetic control analysis using temperature-dependent $R_0$ instead of mean temperature as a covariate.** (A) Shows the total observed cases (black) across all districts with extreme precipitation over time compared to the total cases in the synthetic control (red). (B) Shows the effect of extreme precipitation over time, estimated as the difference between observed cases and synthetic control cases, with the grey ribbon corresponding to the 95% confidence interval. The dashed horizontal line indicates no effect and the dashed vertical line indicates when the cyclone occurred.

#### S1.8 Sensitivity to including observations prior to 2016

We conducted the analysis including observations prior to 2016, starting in 2010. We find that cases were significantly increased by extreme precipitation across a longer period than cases were elevated because of extreme precipitation according to the main analysis (April 22nd - November 3rd). From April 22nd - July 14th, 25,712 (95% CI: 16,320 - 30,978) cases were attributable to extreme precipitation, or 70% (95% CI: 44% - 84%) of cases, an estimate that is slightly larger than that in the main text. There is a slight difference between the synthetic control and observed cases during the 2015 outbreak, although this difference is considerably smaller than that observed during the 2023 outbreak. The synthetic control also regularly exceeds observed outbreaks between 2010 and 2021, suggesting that the extended time series produces a considerably worse control and ultimately less reliable estimates of attributable cases.

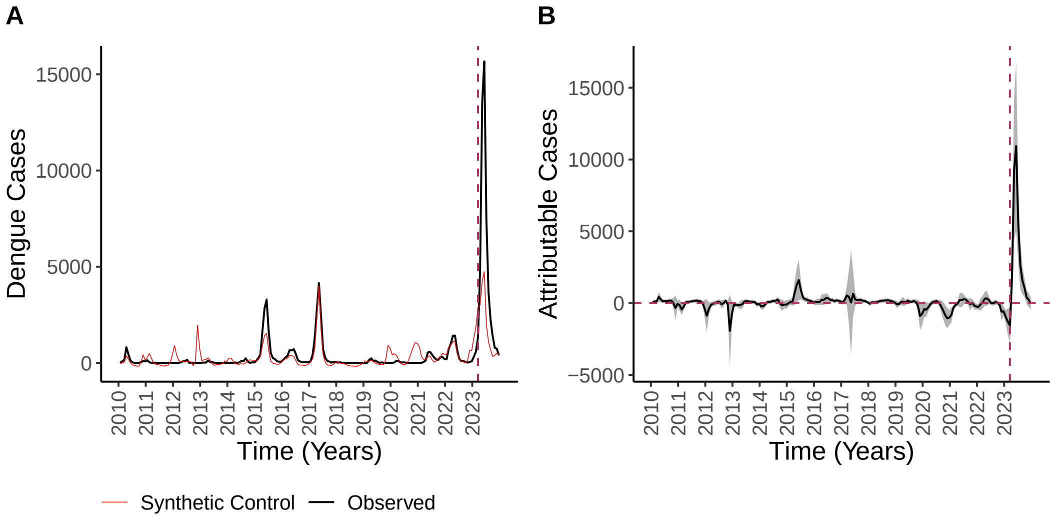

**Figure S15: Results of generalized synthetic control analysis including observations prior to 2016.** (A) Shows the total observed cases (black) across all districts with extreme precipitation over time compared to the total cases in the synthetic control (red). (B) Shows the effect of extreme precipitation over time, estimated as the difference between observed cases and synthetic control cases, with the grey ribbon corresponding to the 95% confidence interval. The dashed horizontal line indicates no effect and the dashed vertical line indicates when the cyclone occurred.

#### S1.9 Sensitivity to excluding observations from 2020 - 2021

We conducted the analysis excluding observations from 2020 - 2021 due to the potential for substantial biases in reporting during the beginning of the COVID-19 pandemic. To conduct this analysis, we additionally included observations from 2010 - 2016 so that a sufficient number of years of observations were used to fit the model. We find that cases were significantly increased by extreme precipitation between March 25th and October 6th, a time period slightly longer than that in the main analysis (Figure S16). Between April 22nd and July 14th, 25,511 (95% CI: 16,705 - 30,531) cases were attributable to extreme precipitation, or 69% (95% CI: 46% - 83%) of all cases.

This estimate is similar to the previous estimate including 2020 - 2021 (subsection S1.8), suggesting that potential biases in case reporting during the COVID-19 pandemic did not substantially affect our results. Cases were generally low during this time period across the districts that experienced extreme precipitation during Cyclone Yaku and variation in dengue cases over time connected to the COVID-19 pandemic that were consistent across the study region were accounted for by the latent factors (Figure 2).

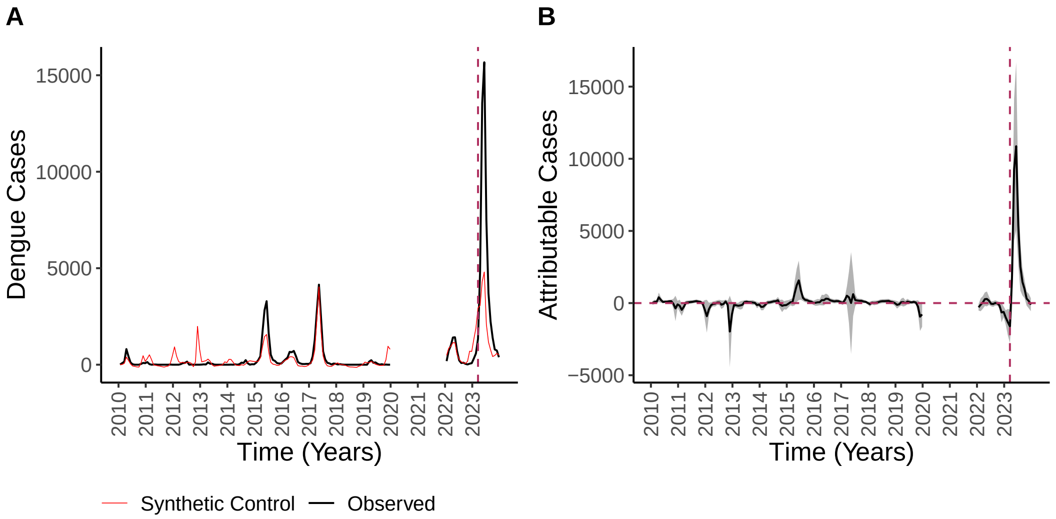

**Figure S16: Results of generalized synthetic control analysis excluding observations from 2020 and 2021.** (A) Shows the total observed cases (black) across all districts with extreme precipitation over time compared to the total cases in the synthetic control (red). (B) Shows the effect of extreme precipitation over time, estimated as the difference between observed cases and synthetic control cases, with the grey ribbon corresponding to the 95% confidence interval. The dashed horizontal line indicates no effect and the dashed vertical line indicates when the cyclone occurred.

#### S1.10 Sensitivity to matching on additional variables

We conducted the analysis with additional matching on the proportion of people residing in land classified as infrastructure by MapBiomas and the population-weighted mean population density (derived from the INEI National Census Raster). This worsened the imbalance between the districts affected by extreme precipitation and the matched control districts; mean precipitation across the matched control districts was 5.4 mm/day (compared to 4.9 mm/day across the matched control districts in the main analysis and 3.7 mm/day in the districts affected by extreme precipitation). Using this matched control pool, we find that extreme precipitation increased dengue cases over a slightly earlier period compared to the main analysis (March 25th - June 16th) (Figure S17). Between April 22nd and July 14th, we estimate that 15,381 (95% CI: -2937 - 25,840) cases were attributable to extreme precipitation or 42% (95% CI: -0.08% - 0.70%) of all cases. The model predictive accuracy was considerably worse compared to the main analysis ($R^2 = 0.25$). Based on this, we conclude that matching on these additional variables worsens balance with respect to precipitation and the fit of the model to case in the districts affected by extreme precipitation.

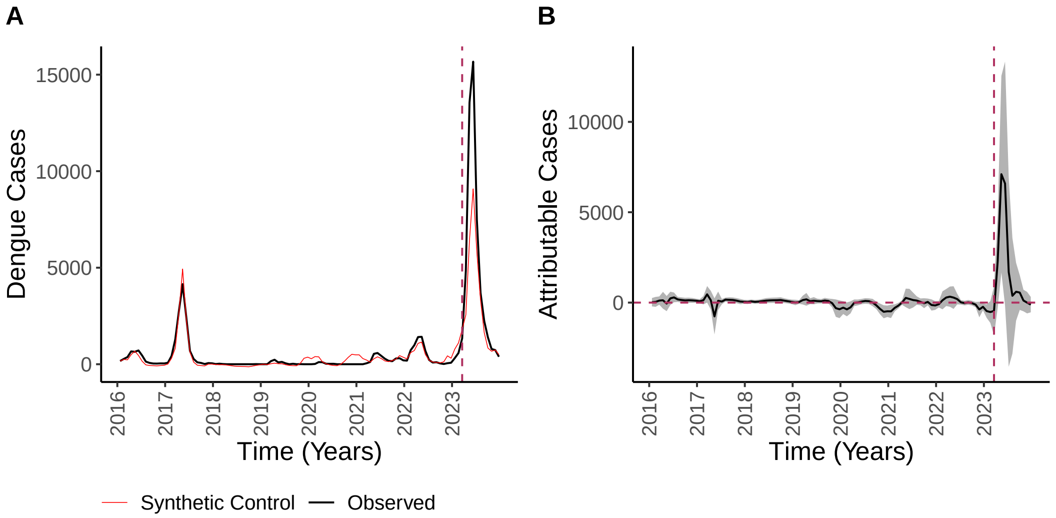

**Figure S17: Results of generalized synthetic control analysis with additional matching on population density and infrastructure.** (A) Shows the total observed cases (black) across all districts with extreme precipitation over time compared to the total cases in the synthetic control (red). (B) Shows the effect of extreme precipitation over time, estimated as the difference between observed cases and synthetic control cases, with the grey ribbon corresponding to the 95% confidence interval. The dashed horizontal line indicates no effect and the dashed vertical line indicates when the cyclone occurred.

##### S1.11 Sensitivity to upper and lower precipitation anomaly thresholds

We repeated the analysis varying the upper precipitation anomaly threshold above which districts were considered to have experience extreme precipitation and the lower precipitation anomaly threshold below which districts were included in the control group with non-extreme precipitation. Note that more districts are classified as experiencing extreme precipitation when the upper threshold is lowered, increasing the number of observed cases under consideration. All numbers and percentages of attributable cases reported here were calculated from April 22nd to July 14th to facilitate comparison with the main model.

When the upper threshold is fixed at 8.5 mm/day and the lower threshold is increased to 8.5 mm/day, the balance with respect to precipitation and temperature improved, while climate covariate balance worsened when the lower threshold was reduced to 5.5 mm/day. In both cases, $R^2$, our measure of model fit (where greater values indicate better fit), decreases compared to the main model (for lower thresholds of 8.5 mm/day: $R^2 = 0.38$; for lower threshold of 5.5 mm/day: $R^2 = 0.13$). The estimate of the percentage of cases attributable to extreme precipitation decreases with a lower threshold of 8.5 mm/day to 28% (95% CI: -31% - 67%) and increases with a lower threshold of 5.5 mm/day to 81% (95% CI: 62% - 91%).

When the upper threshold is increased to 10 mm/day, the percentage of attributable cases is 58% (95% CI: 12% - 87%), 40% (95% CI: -18% - 74%), 22% (95% CI: -54% - 62%), and 7% (95% CI: -83% - 55%) for lower thresholds of 5.5, 7, 8.5, and 10 mm/day, respectively. These estimates are less than those in the main analysis, potentially indicating that greater precipitation anomalies were associated with a smaller increase in cases. $R^2$ was considerably worse that of the main model for a greater upper threshold except when the lower threshold was 5.5 ($R^2 = 0.83$).

The estimate of the percent of cases attributable to extreme precipitation with an upper threshold of 7 mm/day and lower threshold of 7 mm/day is similar to that of the main model: 68% (95% CI: 23% - 83%) of total cases. When the lower threshold is reduce to 5.5 mm/day, we estimate that 82% (95% CI: 23% - 105%) of cases were attributable to the extreme precipitation. The detection of a significant effect of extreme precipitation when the lower threshold is reduced suggests that effects may be observed in districts with precipitation anomalies greater than 5.5 mm/day and that the main analysis therefore underestimates the true number of cases attributable to extreme precipitation for several reasons. First, districts with precipitation anomalies that could lead to increases in cases were excluded from our main analysis. Second, the inclusion of these districts in the control pool may bias our estimate of the cyclone effects downward. However, decreasing the lower and upper thresholds reduces the number of control districts while increasing the number of districts with extreme precipitation, slightly worsening the model fit.

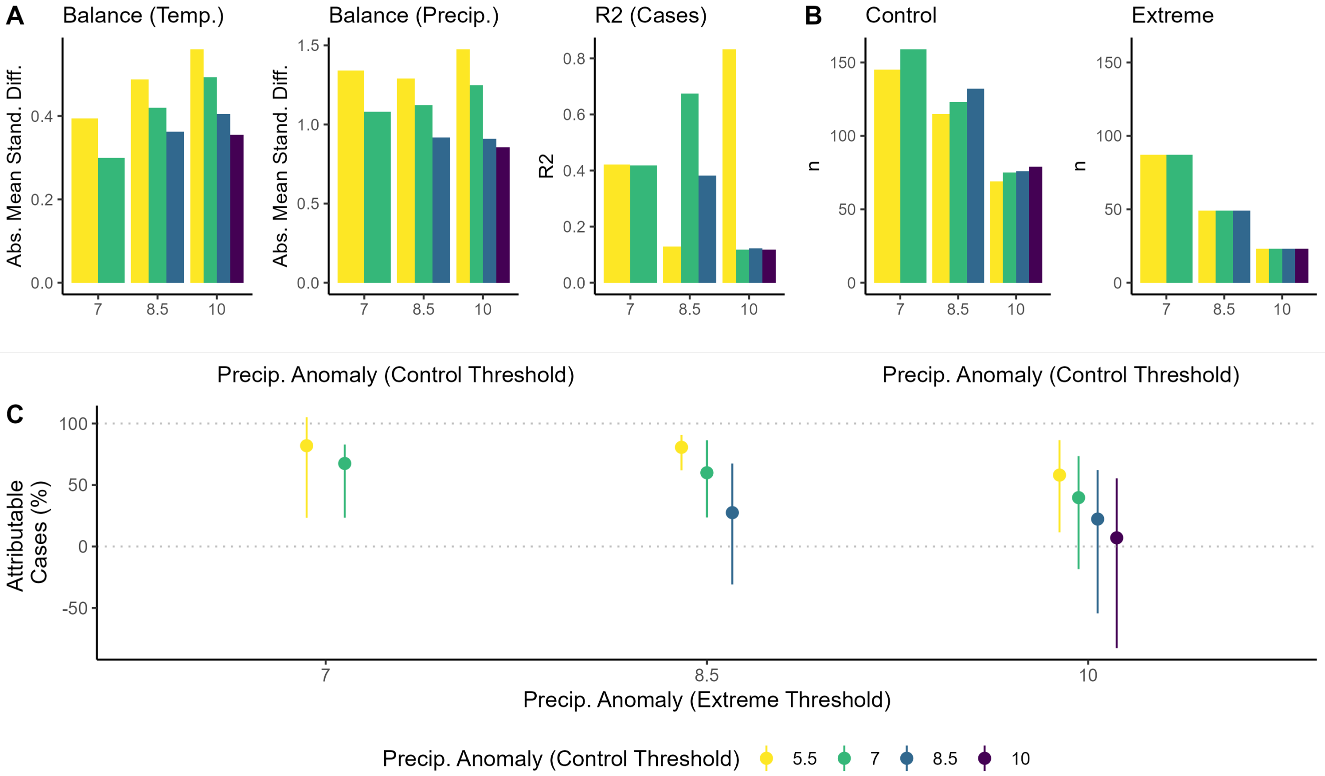

**Figure S18: The effects of varying the upper and lower precipitation anomaly threshold**. The x-axis of each panel is the upper precipitation anomaly threshold. The colors of the bars correspond to the lower precipitation anomaly threshold (legend at bottom). (A) The balance between the districts with extreme precipitation and matched control districts with respect to climate covariates and the prediction error of the resulting generalized synthetic control model depending on upper and lower anomaly thresholds. The first two graphs show balance with respect to temperature (left) and precipitation (middle). Balance is the difference in climate conditions between the districts with extreme precipitation and control districts, measured as the average absolute standardized difference across the study period (Figure S4). The final graph (right) shows $R^2$, a measure of the difference between the observed and predicted (synthetic control) cases prior to the cyclone in the districts that experienced extreme precipitation during Cyclone Yaku. (B) The size of the matched control (left) and extreme precipitation (right) groups. (C) The estimated percentage of cases observed attributable to extreme precipitation across all affected districts between April 22nd and July 14th. The bars indicate 95% confidence intervals.

#### S1.12 Sensitivity to number of matched units

We repeated the main analysis varying the number of control units to which each district with extreme precipitation was matched. The percentage of cases attributable to extreme precipitation increased to 68% (95% CI: 24% - 86%) and decreased to 57% (95% CI: 16% - 81%) when the number of units to match to was increased to ten or fifteen respectively (Figure S19). Model performance was worse for both models ($R^2$ : 0.63 and 0.58 for ten or fifteen matched units respectively). Including all districts with non-extreme precipitation in the control pool for the generalized synthetic control model leads us to estimate that 74% (95% CI: 48% - 81%) of cases were attributable to extreme precipitation, a larger percentage compared to the main analysis, although there is reduced predictive accuracy compared to the main model ($R^2 = 0.51$).

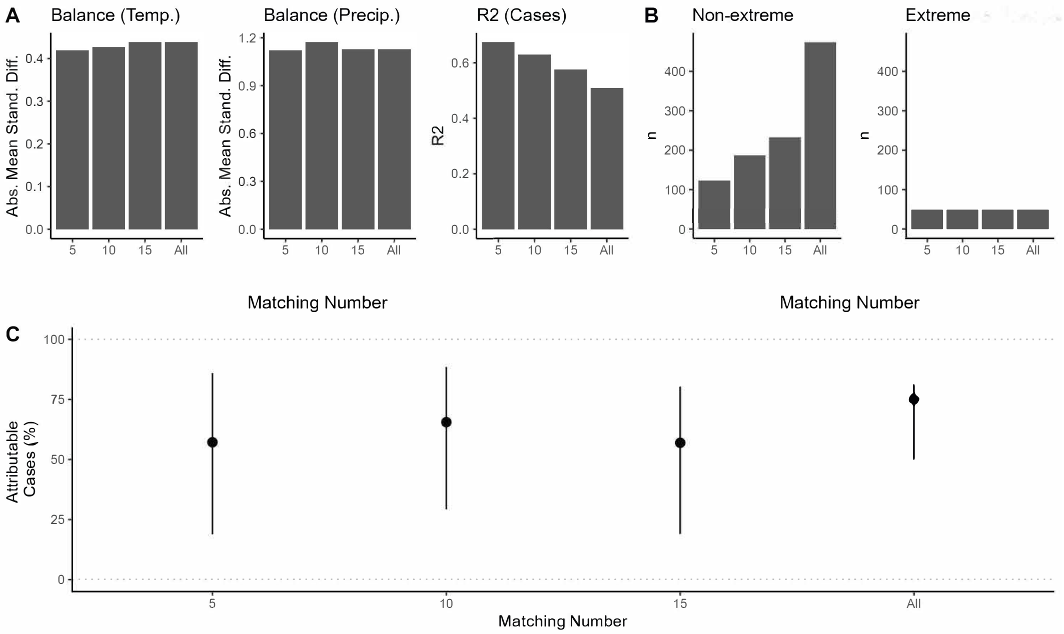

**Figure S19: The effects of varying the number of control districts matched to each district with extreme precipitation (x-axis).** Note that "all" indicates that no matching was performed and all districts with non-extreme precipitation were included in the generalized synthetic control analysis. (A) The balance between the districts with extreme precipitation during Cyclone Yaku and matched control units with respect to climate covariates and the prediction error of the resulting generalized synthetic control model depending on number of matched units. The first two graphs show balance with respect to temperature (left) and precipitation (middle). Balance is measured as the mean absolute value of the standardized difference across the study period (Figure S4). The final graph (right) shows the $R^2$, a measure of the difference between the observed and predicted (synthetic control) cases prior to the cyclone in the districts that experienced extreme precipitation during Cyclone Yaku. (B) The size of the matched control (left) and extreme precipitation (right) groups. (C) The estimated percentage of cases observed attributable to extreme precipitation across all affected districts between April 22nd and July 14th. The bars indicate 95% confidence intervals.

#### S1.13 Sensitivity to number of latent factors

We repeated the main analysis, varying the number of latent factors included in the generalized synthetic control model from zero to five. As described in the main text, the number of latent factors was selected to minimize mean squared prediction error calculated through a cross-validation procedure and five latent factors were therefore included in the main model. The estimated percentage of attributable cases is relatively stable across different numbers of latent factors, ranging from 61% with one latent factor to 76% with four latent factors.

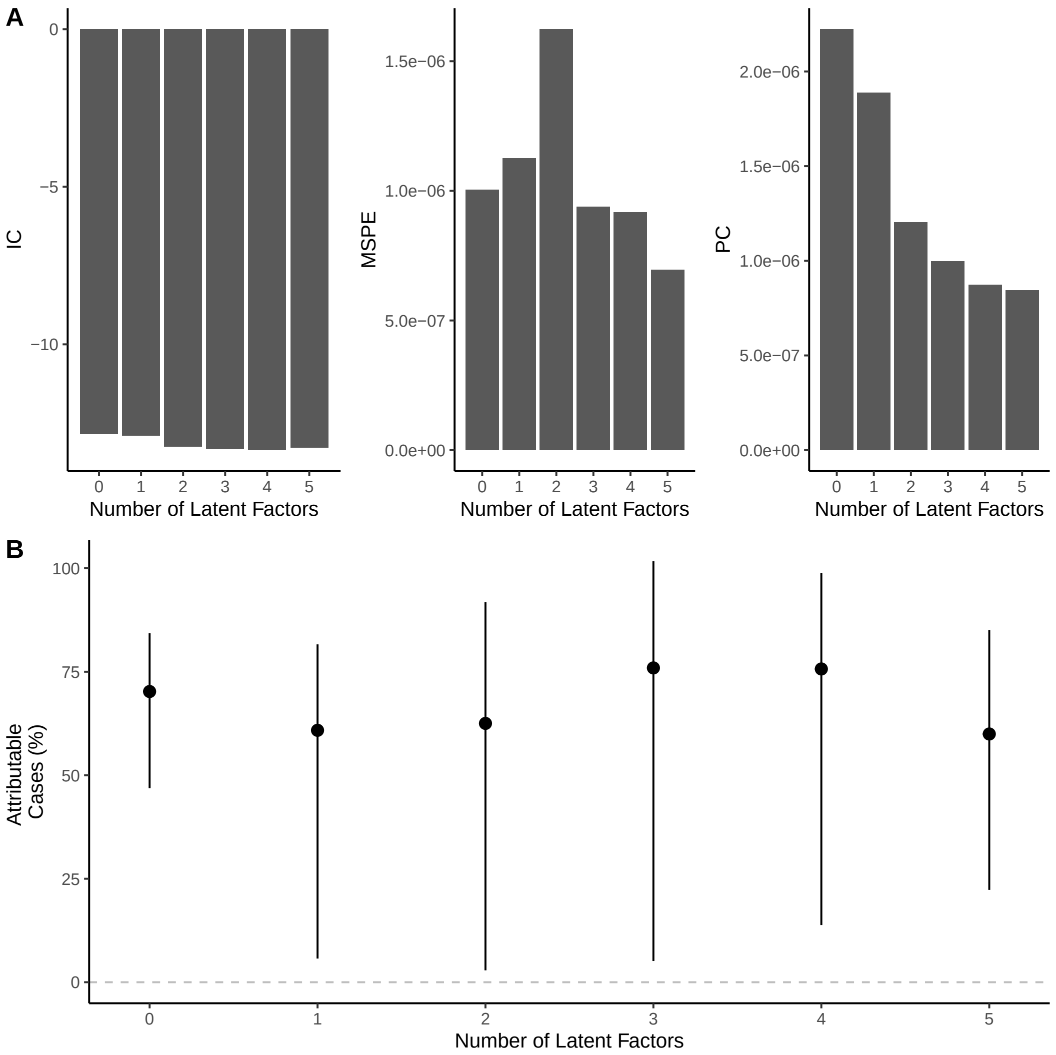

**Figure S20: The effects of varying the number of latent factors (x-axis).** (A) The model fit depending on the number of latent factors. Graphs show the information criterion (IC), proposed criterion (PC), and root mean squared error (RMSE). (B) The estimated proportion of cases that were attributable to extreme precipitation across all affected districts depending on the number of latent factors. The bars indicate 95% confidence intervals.

##### S1.14 Sensitivity to exclusion of Las Piedras

We noted that Las Piedras, Tambopata, Madre de Dios is a matched district with non-extreme precipitation with factor loading values for latent factors 1 and 2 that are especially large in magnitude (-0.007 and 0.003) compared to the mean absolute values across all districts ($2.66 * 10^-4$ and $3.39 * 10^-4$) (Figure S8, Figure S21). We therefore tested the sensitivity of the analysis to excluding this district. As in the main model analysis, we observe that extreme precipitation significantly increased dengue cases from April 22nd - July 14th. Across this period, extreme precipitation caused 22,403 (95% CI: 9,590 - 31,174) cases or 61% (95% CI: 26% - 85%) of all cases.

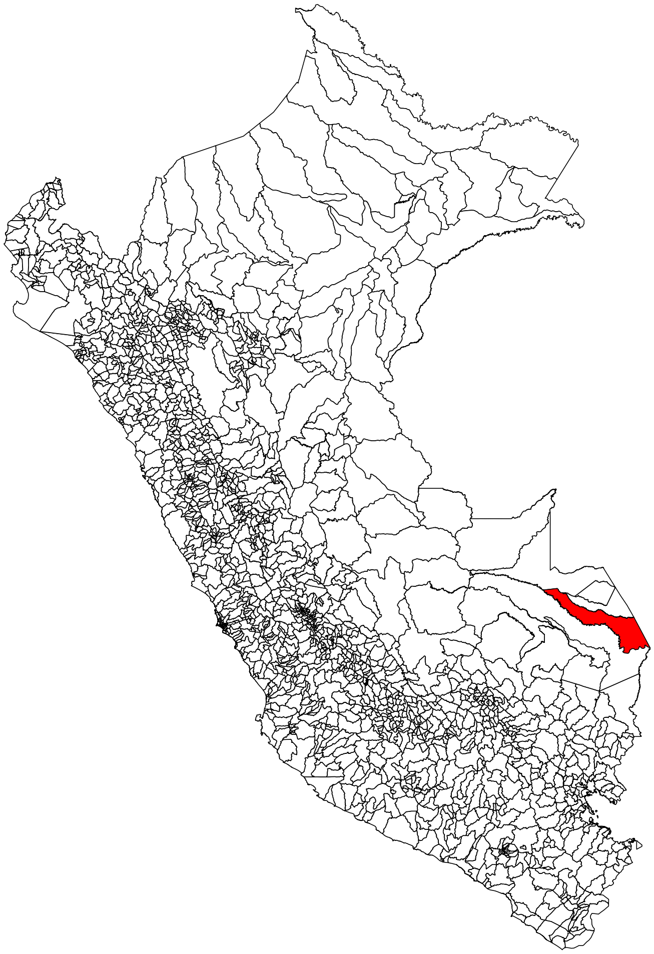

**Figure S21: Map indicating Las Piedras.** A map of Peru with districts outlined in black. Las Piedras, the district excluded from this analysis, is indicated in red.

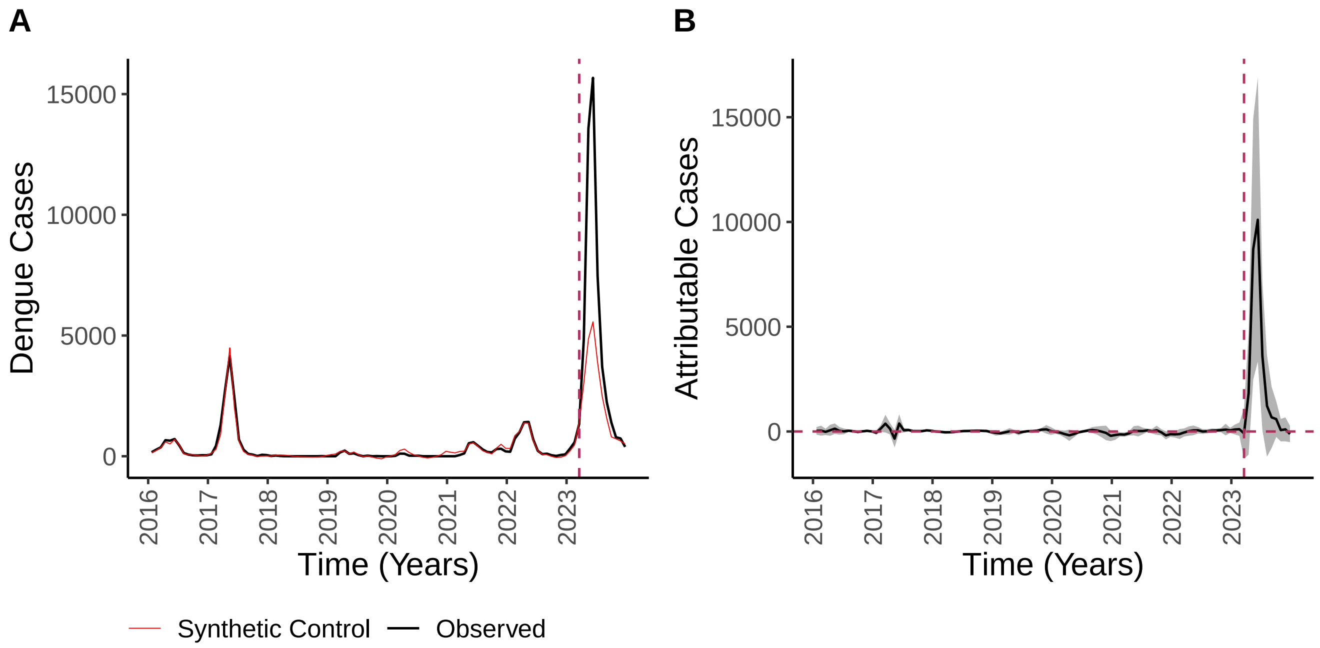

**Figure S22: Results of generalized synthetic control analysis excluding Las Piedras.** (A) Shows the total observed cases (black) across all districts with extreme precipitation over time compared to the total cases in the synthetic control (red). (B) Shows the effect of extreme precipitation over time, estimated as the difference between observed cases and synthetic control cases, with the grey ribbon corresponding to the 95% confidence interval. The dashed horizontal line indicates no effect and the dashed vertical line indicates when the cyclone occurred.

#### S1.15 Supplemental material: examining associations between climate, vulnerability indices, and incidence attributable to extreme precipitation

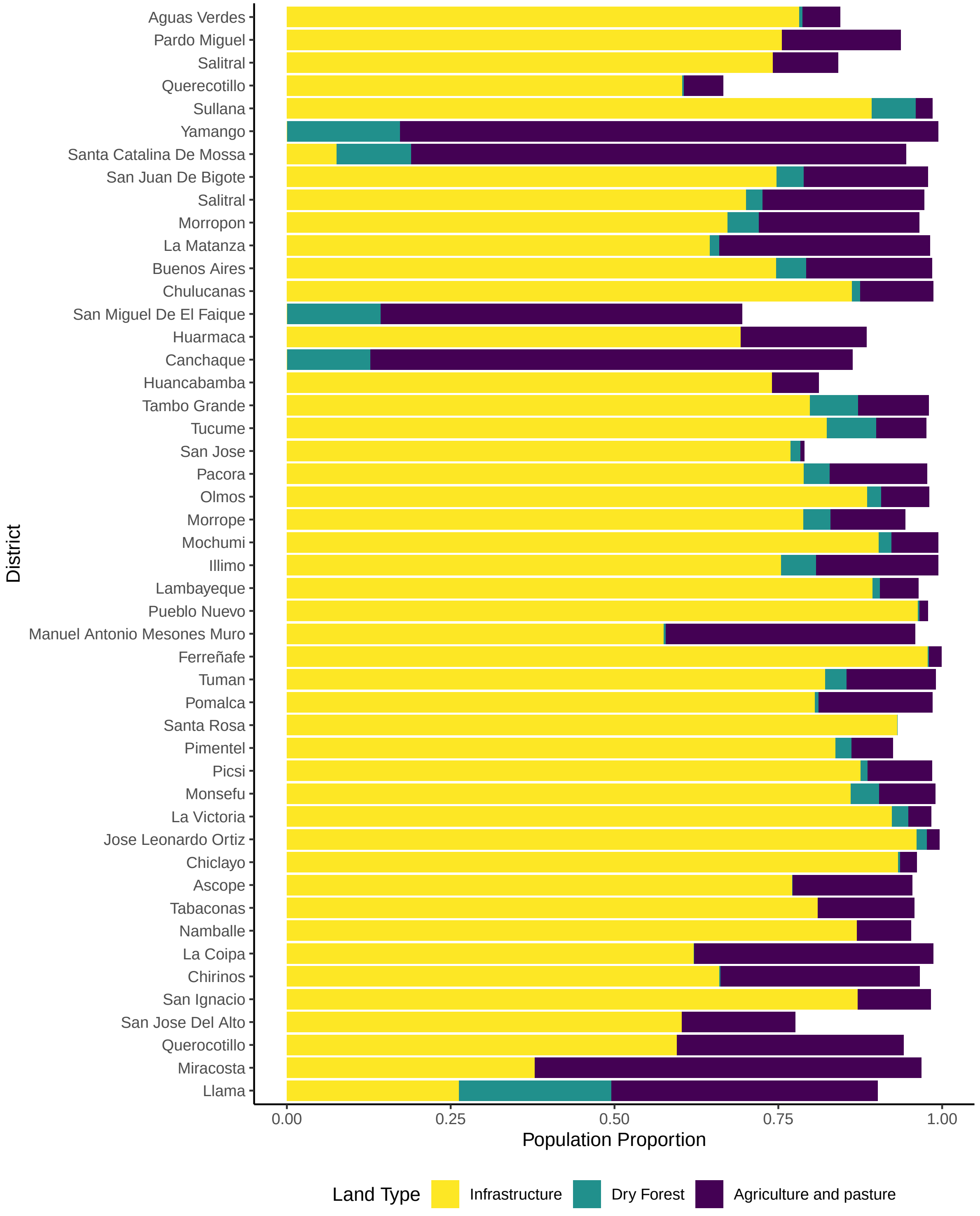

**Figure S23: The predominant land use types in the region affected by extreme precipitation are: infrastructure, agriculture and pasture, and dry forest).** Stacked barplot displaying the proportion of the population residing in land of a given type (y-axis) across districts (x-axis). The land use types included are infrastructure (yellow), dry forest (green), and agriculture and pasture (purple).

| Name | Description | Source |
|---|---|---|
| Distance to roads (meters) | 50 m resolution raster. Continuous variable. If less than 500 m from the nearest road, 1. If greater than 3000, 0. | MTC. 2020. |
| Distance to natural bodies of water and rivers (meters) | 50 m resolution raster. Continuous variable. If less than 500 m from the nearest body of water, 1. If greater than 1500, 0. | MTC. 2023. |
| Non-public water source | Manzana-level. Proportion of residences not connected to the public water network. | INEI. 2017 National Census. |
| Inconsistent water access | Manzana-level. Proportion of residences without access to water two or more days per week. | INEI. 2017 National Census. |
| Household overcrowding | Manzana-level. Proportion of residences with overcrowding (i.e., the number of people to rooms in a dwelling is greater than or equal to 3). | INEI. 2017 National Census. |
| Low-quality residences | Manzana-level. Proportion of residences that were low-quality (e.g., hut or cabin, improvised residence, premises not intended for human residents). | INEI. 2017 National Census. |
| Susceptibility to flooding | 200 m resolution raster. Categorical variable. Forecast for Jan - March 2024. Based on geomorphology and terrain slope. Values can be: very low (0), low (1), medium (2), high (3), very (high). | INGEMETT, CENEPRED (CENEPRED 2023). 2023. |
| Infrastructure | 30 m resolution. Continuous variable. Proportion of population living in land classified as infrastructure. | MapBiomas, derived from Landsat images (Instituto del Bien Común (IBC) 2022). 2022. |
| Agriculture and pasture | 30 m resolution. Continuous variable. Proportion of population living in land classified as agriculture, pasture, or agriculture and pasture mosaic. | MapBiomas, derived from Landsat images (Instituto del Bien Común (IBC) 2022). 2022. |
| Dry forest | 30 m resolution. Continuous variable. Proportion of population living in land classified as dry forest. | MapBiomas, derived from Landsat images (Instituto del Bien Común (IBC) 2022). 2022. |

**Table S5: Vulnerability indices.** Each row corresponds to a different vulnerability index, provided by CDC Peru. The first column gives the names of the indices. The second gives descriptions of the indices, including units, resolution, and information on how they were calculated. The third column gives the abbreviated name of the Peruvian agency that provided data for a given index and the year for which it was defined (for the final row, flood risk in 2024 was calculated based on 2023 data). Agency names are abbreviated as following: MTC (Ministerio de Transportes y Comunicaciones, Minister of Transport and Communications); INEI (Instituto Nacional de Estadistica e Informatica, National Institute of Statistcs and Information); INGEMETT (Instituto Geológico, Minero y Metalúrgico; The Geological Mining and Metallurgical Institute; CENEPRED (Centro Nacional de Estimación Prevención y Reducción del Riesgo de Desastres, The National Center for Estimation, Prevention and Reduction of Disaster Risk).

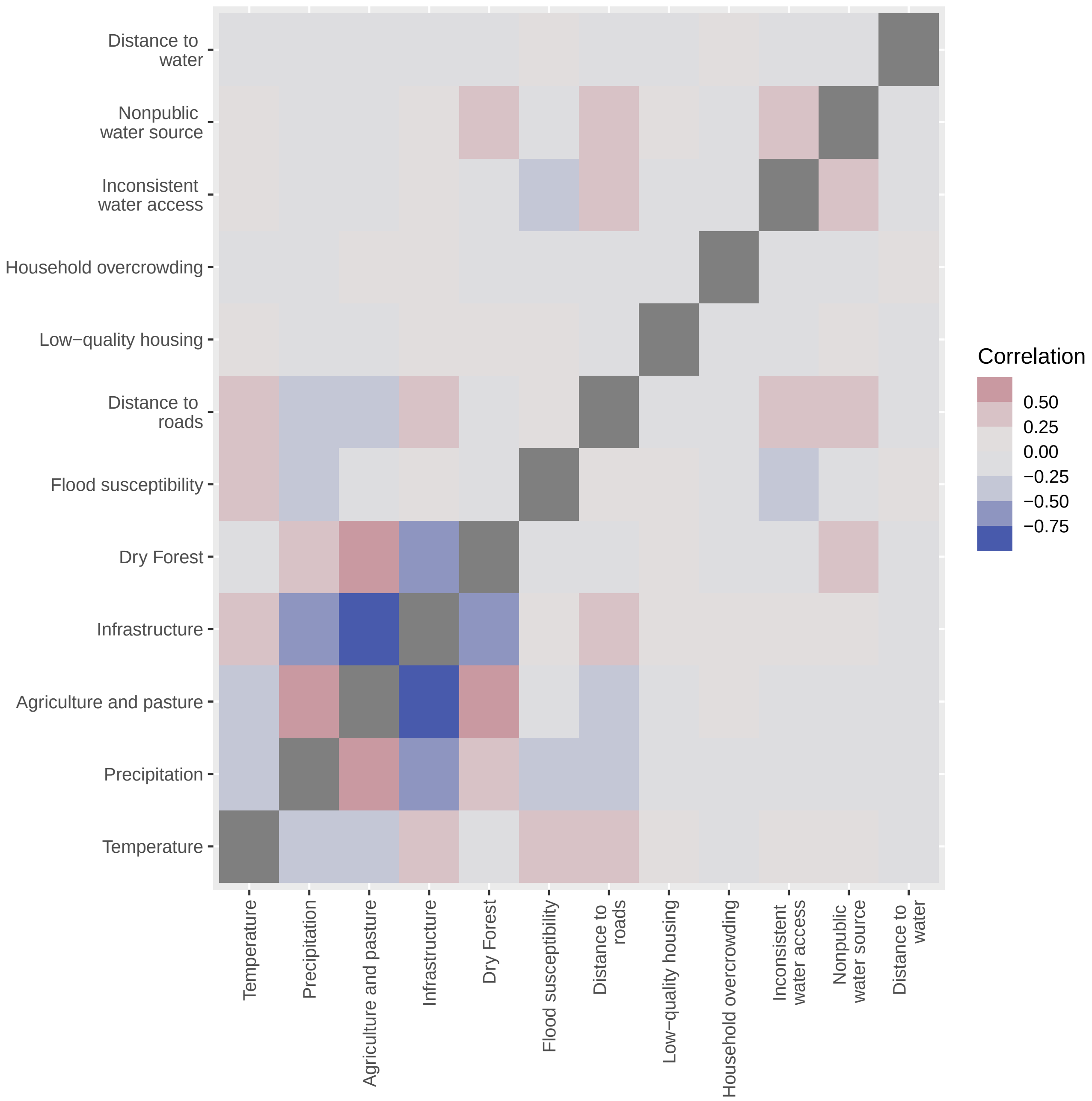

**Figure S24: Vulnerability indices were correlated with each other.** Heatmap displaying the correlation coefficient between different vulnerability indices (labeled along x- and y-axes, see Table S5). More positive correlations are indicated in darker shades of red and more negative correlations are indicated in darker shades of blue. The diagonal (which would display a perfect correlation between each variable and itself) is indicated in grey.

| | RC1 | RC2 | RC3 |
|---|---|---|---|
| Infrastructure | **0.94** | -0.03 | 0.04 |
| Distance to roads | 0.52 | 0.52 | 0.18 |
| Temperature | 0.51 | 0.29 | 0.39 |
| Inconsistent water access | 0.25 | **0.63** | -0.34 |
| Flood susceptibility | 0.23 | -0.34 | **0.76** |
| Distance to body of water | 0.16 | -0.49 | -0.14 |
| Nonpublic water source | 0.09 | **0.80** | -0.06 |
| Household overcrowding | 0.03 | -0.17 | -0.57 |
| Low-quality housing | 0.00 | -0.01 | 0.48 |
| Dry Forest | **-0.73** | 0.43 | 0.24 |
| Precipitation | **-0.77** | -0.08 | -0.07 |
| Agriculture and pasture | **-0.92** | -0.11 | -0.01 |

**Table S6: Factor loadings for each rotated component.** Across each row, we display the standardized loadings for each vulnerability index (see Table S5) with respect to rotated components RC1, RC2, and RC3. Rows are ordered in descending order based on the loading for RC1. Bolded values indicate loadings with absolute value greater than 0.5.

| Rotated Component | Coefficient | p-value |
|---|---|---|
| Infrastructure (+),<br>Agriculture and Pasture (-) ($RC_1$) | 5.20 (2.72 - 8.05) | 0.000 |
| Nonpublic water source (+),<br>Inconsistent water access (+) ($RC_2$) | -1.20 (-4.25 - 1.76) | 0.232 |
| Flood susceptibility (+),<br>Household overcrowding (-) ($RC_3$) | 6.11 (3.43 - 8.71) | 0.000 |
| Y-intercept | 4.05 (2.70 - 7.80) | 0.014 |

**Table S7: Values estimated for coefficients of the rotated components (derived from vulnerability indices and climate) and y-intercept.** The factors most strongly associated with each rotated components are listed in the first column with their direction indicated in parentheses as either positive (+) or negative (-) (see Table S6). We additionally provide bootstrapped 95% confidence intervals and p-values for estimates.

##### S1.16 Supplemental material: quantifying the influence of historical climate forcing on the probability of extreme March precipitation in northwestern Peru

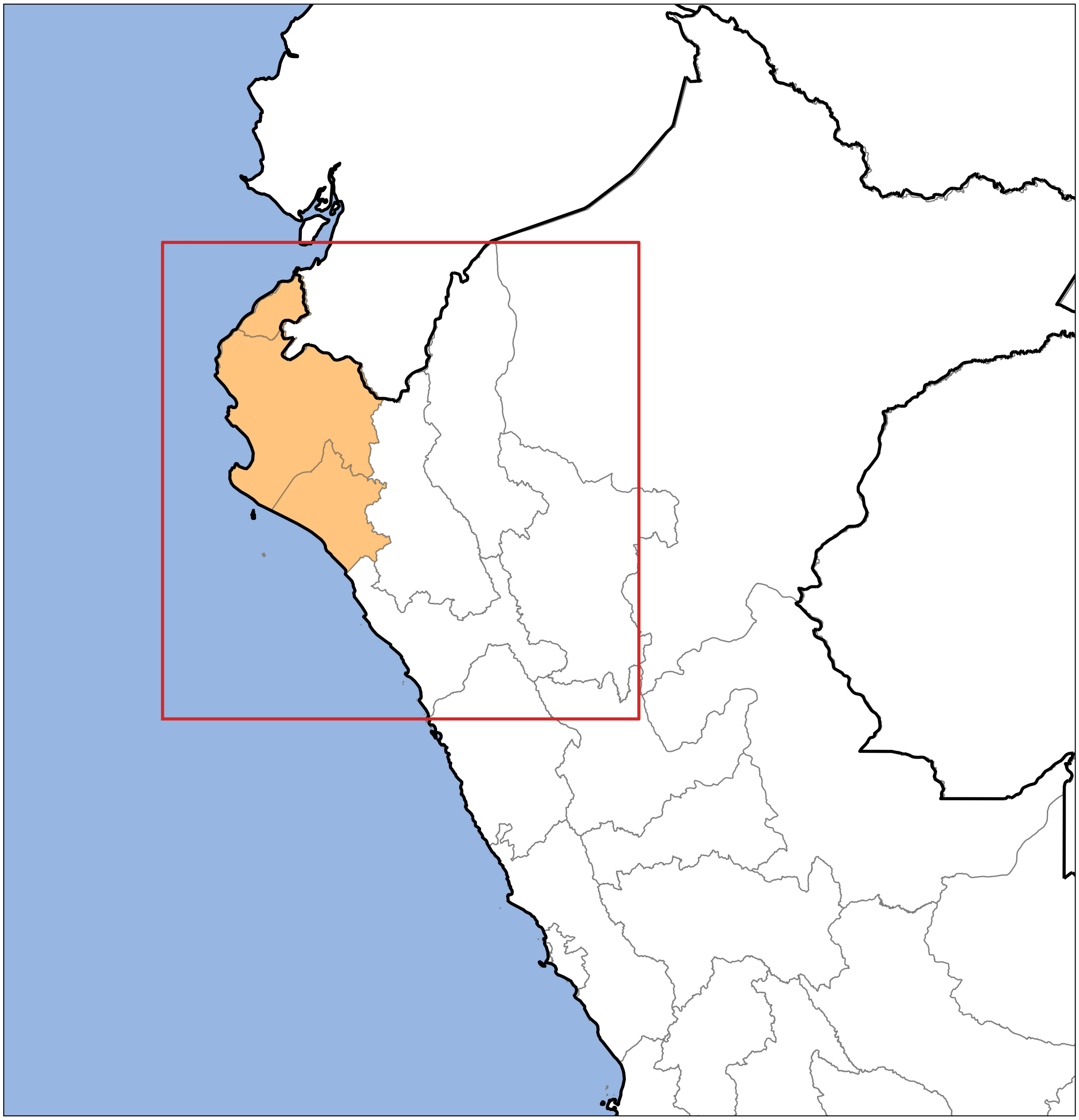

**Figure S25: Map of the region of Northwestern Peru used in the climate attribution analysis.** A map of Peru is displayed with national borders indicated with thick lines and regional borders indicated with thin lines. The red box indicates the region of northwestern Peru used in the precipitation attribution analysis. The orange shaded region indicates the three northwestern coastal districts (Tumbes, Piura, and Lambayeque) used in the temperature attribution analysis.

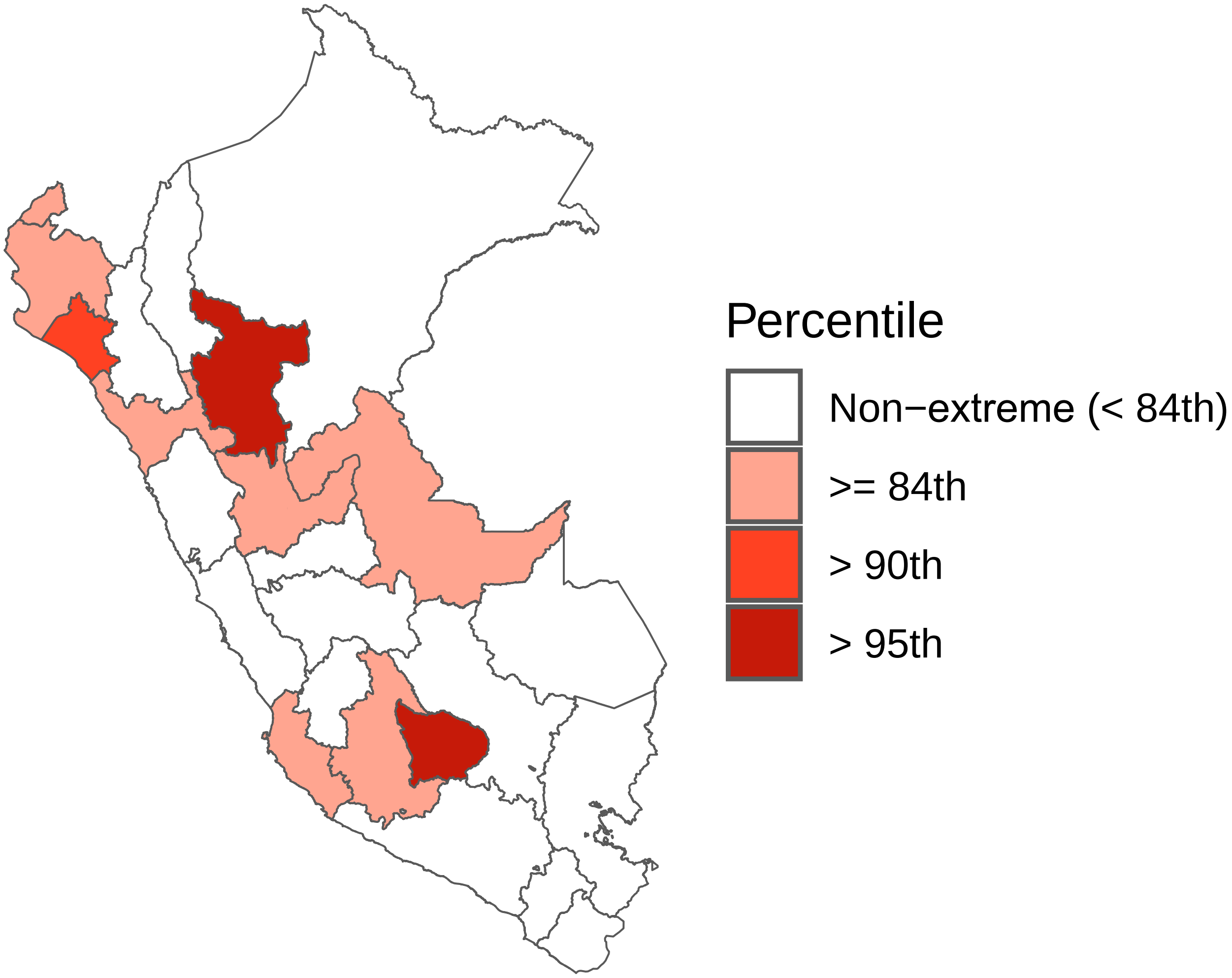

**Figure S26: Regions with extreme precipitation exceeded the 84th percentile for precipitation during Cyclone Yaku.** Precipitation was defined as the average population-weighted total precipitation for each region in the month of March. Precipitation in 2023 (i.e., during Cyclone Yaku) was compared to observations from 1973 - 2022. White indicates regions where precipitation did not exceed the 84th percentile during Cyclone Yaku (i.e., non-extreme precipitation), whereas shades of red correspond to precipitation above the 84th percentile, where darker shades of red indicate more extreme precipitation.

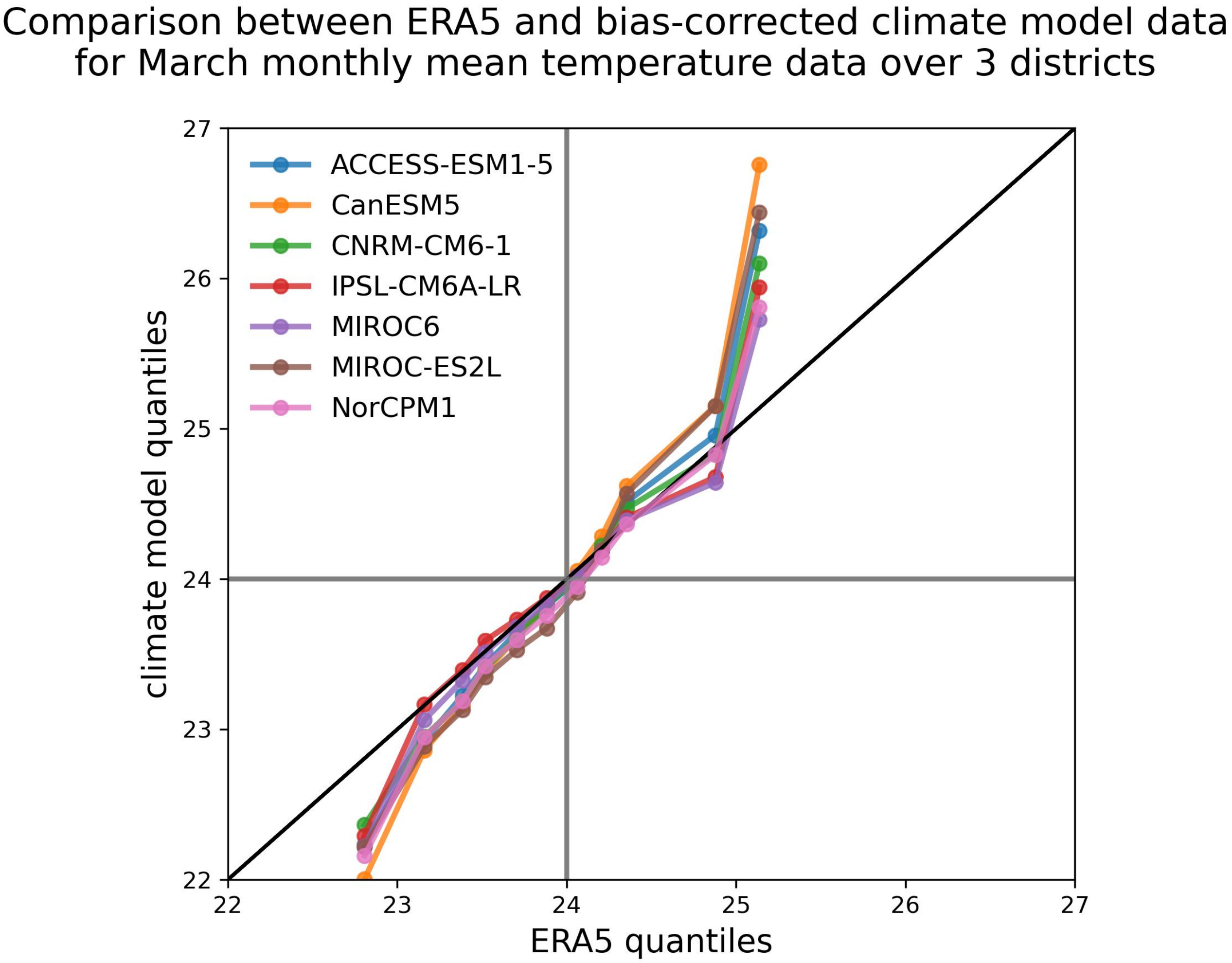

**Figure S27: Quantile-quantile plot comparing ERA5 and bias-corrected climate model data for the 1985-2014 period.** Comparison between March monthly mean temperature quantiles averaged over three northwestern coastal districts (Lambayeque, Piura, and Tumbes) in ERA5 reanalysis data and debiased climate model data which has been downscaled to match the ERA5 spatial resolution using the bias-correction and spatial disaggregation approach (see Methods).

#### S1.17 Sensitivity to using a percentile-based threshold for suitably warm temperature

We repeated the climate attribution analysis using a percentile-based threshold for suitably warm temperature instead of an absolute threshold of $24°C$. We determined that $24°C$ corresponds to the 54th percentile for the recent climate (Figure S28). Using this definition, the frequency of suitably warm temperature has increased from 12.65% during the pre-industrial period to 20.91% in the early historical period and 46.00% in the late historical period (Figure S29). Since the pre-industrial era, the likelihood of suitably warm temperature increased by 65% in the early historical period and 264% in the late historical period. Between the early and late historical period, the likelihood of suitably warm temperature increased by 120%. The likelihood of co-occurring of extreme precipitation and suitably warm temperature increased from 3.71% to 5.68% to 10.97% in the preindustrial, early historical, and late historical periods respectively. Compared to the preindustrial period, extreme precipitation and suitably warm temperature were 53% more likely and 196% more likely to co-occur in the early and late historical periods respectively; co-occurence was 93% more likely in the late historical period compared to the early historical period. All increases were statistically significantly ($p < 0.001$), as in the main analysis.

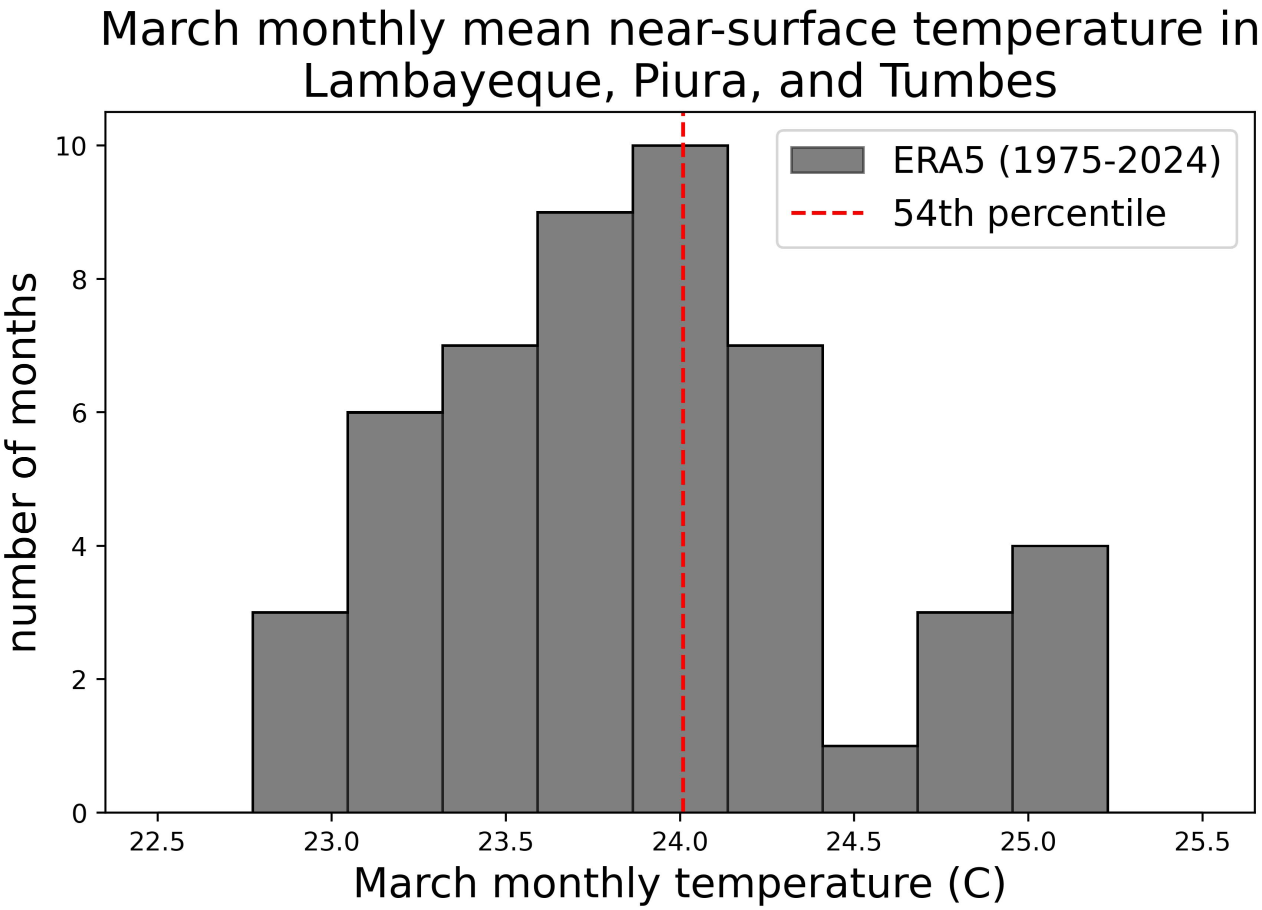

**Figure S28: The 54th percentile for mean temperature corresponded to $24°C$ in three coastal regions of northwestern Peru that experienced extreme precipitation in March 2023.** We plot the distribution of the mean temperature (from ERA5 data) across Lambayeque, Piura, and Tumbes in the month of March across the years from 1975-2024. The red dashed line indicates that $24°C$ corresponds to the 54th percentile across the distribution.

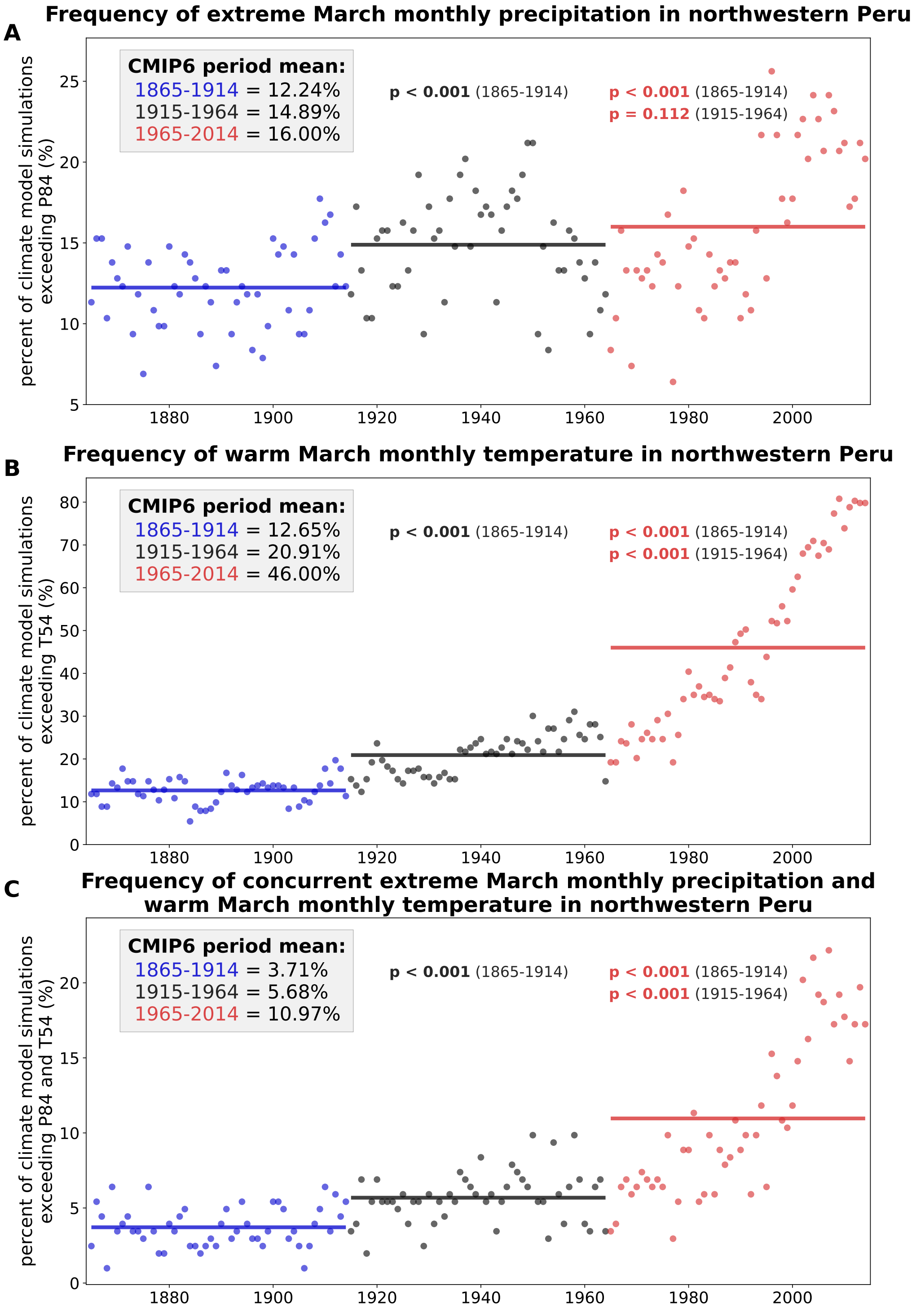

**Figure S29: Climate attribution analysis results are robust to defining suitably warm temperature using a percentile-based threshold.** As in Figure 5, each panel corresponds to a different weather condition: (A) extreme precipitation (B) suitably warm temperature and (C) concurrent extreme precipitation and warm temperature. Annual values show the percentage of 203 climate model simulations exhibiting a given climate condition during each calendar year of the CMIP6 historical forcing experiment for the 1865-2014 period. In contrast to the main analysis, suitably warm temperature is defined to have occured when March monthly mean temperature averaged over northwestern Peru exceeds the 54th percentile for the recent climate (calculated from the last 50 years of the CMIP6 historical forcing experiment, or 1965-2014) based on the quantiles observed in 2023 (Figure S28). As in the main analysis, the threshold for extreme precipitation is the 84th percentile (P84) (Figure S26). Solid lines show the mean frequency for the preindustrial (1865-1914, blue), early historical (1915-1964, gray), and late historical (1965-2014, red) periods. We also show p-values for the Kolmogorov-Smirnov (KS) test for the mid- and late-periods calculated relative to the time periods shown in parentheses.

##### S1.18 Debiasing ERA5 temperature data

We compared remotely sensed hourly temperature from ERA5 to monthly average temperatures between 1970 and 2000 reported by the high resolution climatology WorldClim (Fick and Hijmans 2017) following Childs et al. (2024), using the equation:

$$\widehat{ERA5}_{ihmy} = ERA5_{ihmy} - \overline{ERA5_{im}} + \overline{WorldClim_{im}}$$

$\widehat{ERA5}_{ihmy}$ is the debiased ERA5 hourly temperature in a given district (where subscripts i, h, m, and y designate the district, hour, month, and year, respectively), $ERA5_{ihmy}$ is the raw ERA5 hourly temperature in the corresponding district, $\overline{ERA5_{im}}$ and $\overline{WorldClim_{im}}$ are the mean monthly temperatures for a given district from 1970 - 2000 from ERA5 and WorldClim, respectively.
